# Precision medicine in type 2 diabetes: targeting SGLT2 inhibitor treatment for kidney protection

**DOI:** 10.1101/2024.09.01.24312905

**Authors:** Thijs T. Jansz, Katherine G. Young, Rhian Hopkins, Andrew P. McGovern, Beverley M. Shields, Andrew T. Hattersley, Angus G. Jones, Ewan R. Pearson, Coralie Bingham, Richard A. Oram, John M. Dennis, the MASTERMIND Consortium

## Abstract

**Aims/hypothesis:** Current guidelines recommend use of sodium–glucose cotransporter-2 inhibitors (SGLT2 inhibitors) for kidney protection in people with type 2 diabetes and early-stage chronic kidney disease (CKD) based on a urinary albumin/creatinine ratio (uACR) of ≥3 mg/mmol. However, individuals with a normal uACR or low-level albuminuria were not represented in kidney outcome trials, leaving uncertainty about absolute treatment benefit in this group. To address this gap and support treatment decisions in clinical practice, we developed and validated a model to predict individual-level kidney protection benefit through use of SGLT2 inhibitors.

**Methods:** This observational cohort study used electronic health record data from UK primary care (Clinical Practice Research Datalink, 2013–2020) of adults with type 2 diabetes, eGFR ≥60 ml/min per 1.73 m^2^ and uACR <30 mg/mmol, without heart failure or atherosclerotic vascular disease, who were starting treatment with either SGLT2 inhibitors or the comparator drugs dipeptidyl peptidase-4 (DPP4) inhibitors/sulfonylureas. First, we confirmed the real-world applicability of the relative treatment effect from a previous SGLT2 inhibitor trial meta-analysis, using overlap-weighted Cox proportional hazards models. Second, we assessed calibration of the CKD-PC risk score for kidney disease progression (≥50% eGFR decline, end-stage kidney disease or kidney-related death). Third, we integrated the relative treatment effect with the risk score to predict 3-year individual-level absolute risk reductions for SGLT2 inhibitors, and validated the accuracy of predictions vs overlap-weighted estimates based on observed data. Finally, we compared the clinical utility of a model-based treatment strategy with that of the ≥3 mg/mmol albuminuria threshold.

**Results:** In 53,096 initiations of SGLT2 inhibitor treatment compared with 88,404 initiations of DPP4 inhibitor/sulfonylurea treatment, there was a 42% lower relative risk of kidney disease progression with SGLT2 inhibitors (HR 0.58; 95% CI 0.48, 0.69), consistent with a previous trial meta-analysis. The CKD-PC risk score did not require recalibration (calibration slope 1.05; 95% CI 0.94, 1.17). The median overall model-predicted absolute risk reduction with SGLT2 inhibitors was 0.37% at 3 years (IQR 0.26–0.55), and showed good calibration (calibration slope 1.10; 95% CI 1.09, 1.12). As an illustration of clinical utility, using the model predictions to target the same proportion of the population (*n*=25,303, 17.9%) as the albuminuria threshold would prevent over 10% more events over 3 years (253 vs 228) by identifying a subgroup of 6.7% of individuals with uACR <3 mg/mmol who showed significantly greater absolute risk reduction in response to SGLT2 inhibitor treatment than the remainder with uACR <3 mg/mmol (3.2% vs 1.2% in extended 5-year observational analyses, *p*=0.05).

**Conclusions/interpretation:** A model adapting the international CKD-PC risk score can accurately predict the individual-level kidney protection benefit from treatment with SGLT2 inhibitors in people with type 2 diabetes and no or early-stage CKD. This could guide treatment decisions in clinical practice worldwide. and could target treatment more effectively than the ≥3 mg/mmol albuminuria threshold recommended by current international guidelines.

**Research in context:** *What is already known about this subject?:*

- Sodium–glucose cotransporter-2 (SGLT2) inhibitors reduce the risk of kidney failure in people with type 2 diabetes
- Current guidelines recommend use of SGLT2 inhibitors for kidney protection in individuals with type 2 diabetes and urinary albumin/creatinine ratio ≥3 mg/mmol, but this is an extrapolation beyond current evidence from kidney outcome trials
- It is unclear which people with type 2 diabetes and preserved eGFR and a normal urinary albumin/creatinine ratio or low-level albuminuria have clinically relevant kidney protection benefit from SGLT2 inhibitors

*What is the key question?:*

- Can a model integrating an established risk score with the relative treatment effect from a SGLT2 inhibitor trial meta-analysis accurately predict individual-level kidney protection benefit?

*What are the new findings?:*

- The model accurately predicted individual-level kidney protection benefit with SGLT2 inhibitor treatment in an external validation using UK primary care data
- Compared with the ≥3 mg/mmol albuminuria threshold, the model more effectively identified individuals who are likely to benefit, and could prevent more adverse kidney events

*How might this impact on clinical practice in the foreseeable future?:*

- The model enables individualised prescribing of SGLT2 inhibitors for kidney protection, which could optimise treatment allocation and improve kidney outcomes

## Introduction

Sodium–glucose cotransporter 2 (SGLT2) inhibitors are now widely recognised for their effect on lowering risk of kidney failure [1]. Consequently, international guidelines recommend use of SGLT2 inhibitors in individuals with type 2 diabetes, an eGFR ≥20 ml/min per 1.73 m^2^ and chronic kidney disease (CKD): i.e. a reduced eGFR (<60 ml/min per 1.73 m^2^) or albuminuria (urinary albumin/creatinine ratio [uACR] ≥3 mg/mmol) [2, 3]. However, these recommendations are extrapolations beyond existing trial evidence. Notably, SGLT2 inhibitor kidney outcome trials only included individuals with markedly reduced eGFR (20–45 ml/min per 1.73 m^2^) or severe albuminuria (uACR ≥22.6 mg/mmol) and did not include those with preserved eGFR (≥60 ml/min per 1.73 m^2^) and only low-level albuminuria [4–6]. While data from cardiovascular outcome trials suggest that these individuals have similar relative risk reductions for adverse kidney outcomes when treated with SGLT2 inhibitors [7–9], it remains unclear whether they have clinically meaningful absolute risk reductions. Furthermore, this uncertainty applies to individuals with preserved eGFR and normal uACR (<3 mg/mmol), whom current guidelines do not recommend treating with SGLT2 inhibitors.

A more precise alternative to the ≥3 mg/mmol albuminuria threshold could be to target SGLT2 inhibitor treatment based on an individual’s predicted absolute risk reduction. This could be feasible using existing clinical prediction models, such as the internationally validated CKD Prognosis Consortium (CKD-PC) risk score [10] which predicts an individual’s 3-year absolute risk of kidney disease progression using routine clinical features. By integrating this risk score with the relative treatment effect from an SGLT2 inhibitor trial meta-analysis, which trial [7–9] and real-world evidence [11] indicates is consistent across eGFR and albuminuria levels, an individual’s likely absolute risk reduction with SGLT2 inhibitor treatment, i.e. benefit, could potentially be estimated. Similar approaches are widely used in other fields (e.g. for individualised prescribing of statins for cardiovascular protection [12, 13]), but have not yet been applied to kidney protection.

We therefore aimed to develop and validate a risk score-based model to predict absolute risk reductions in kidney disease progression when treated with SGLT2 inhibitors in a lower-risk population with no established CVD and no or early-stage CKD (preserved eGFR and normal uACR/low-level albuminuria). This group represents approximately 75% of people with type 2 diabetes [14]. To achieve this, we first confirmed that the relative treatment effect from SGLT2 inhibitor trial meta-analysis was applicable in this population using real-world primary care data, including across albuminuria levels. Second, we confirmed satisfactory performance of the CKD-PC risk score. We then integrated these two components to predict individual-level absolute risk reductions (pARR) and validated the predictions. Finally, we compared the clinical utility of treatment strategies based on *pARR* with use of the currently recommended albuminuria threshold for SGLT2 inhibitor initiation.

## Methods

### Data sources

This observational cohort study used electronic health record data from the UK-representative Clinical Practice Research Datalink (CPRD) Aurum dataset [15]. CPRD contains information recorded during routine primary care, including demographic characteristics, diagnoses, prescriptions, laboratory tests and physiological measurements. The CPRD data were linked to national hospital inpatient data (Hospital Episode Statistics), Office for National Statistics death data and Index of Multiple Deprivation (IMD) 2015 data.

### Study population

We included individuals with type 2 diabetes who started treatment with SGLT2 inhibitors as well as individuals who were not on SGLT2 inhibitors but started treatment with dipeptidyl peptidase-4 (DPP4) inhibitors or sulfonylureas as comparator drugs (the three most commonly prescribed oral glucose-lowering treatments after metformin [16]) between 21 January 2013 and 15 October 2020, for whom at least 91 days of registration data were available before first prescription. Neither DPP4 inhibitors nor sulfonylureas are known to alter kidney disease progression [17, 18]. Treatment initiation and study baseline were defined the date of a first-ever prescription for an SGLT2 inhibitor or DPP4 inhibitor/sulfonylurea.

We excluded individuals according to the following criteria: (1) a prior diagnosis of heart failure or atherosclerotic CVD (ischaemic heart disease or angina, peripheral vascular disease, revascularisation, stroke or transient ischaemic attack) given the clear indication for treatment with SGLT2 inhibitors due to their cardiovascular benefit [19]; (2) missing baseline eGFR or uACR; (3) eGFR <60 ml/min per 1.73 m^2^ or uACR ≥30 mg/mmol; (4) end-stage kidney disease (ESKD); or (5) concurrent prescription of glucagon-like peptide-1 receptor agonists, due to their kidney-protective effects [20]. To align with the population used for development of the CKD-PC clinical prediction model [10], we also excluded those with BMI <20 kg/m^2^ or age <20 or >80 years.

### Treatment

DPP4 inhibitors and sulfonylureas were considered a single comparator arm due to their similar risk profiles for kidney disease progression [17, 18]. We confirmed the validity of this approach using sensitivity analyses. All individuals were followed from drug initiation until the earliest of the following: outcome event, death, primary care practice deregistration, initiation of treatment with glucagon-like peptide 1 receptor agonists, the last date of the study period, or a maximum of 3 years (with additional analyses extended to 5 years), regardless of whether repeat prescriptions were issued. Individuals in the comparator arm who started treatment with an SGLT2 inhibitor during follow-up were censored at the date of SGLT2 inhibitor initiation. They were subsequently included in the SGLT2 inhibitor arm from the same date, allowing inclusion of two non-overlapping follow-up periods for these individuals. We did not censor individuals in either arm who started treatment with DPP4 inhibitors or sulfonylureas during follow-up.

### Outcomes

The primary outcome was kidney disease progression, defined as a composite of a ≥50% decline in eGFR, ESKD (requirement for renal replacement therapy or sustained eGFR <15 ml/min per 1.73 m^2^) or death from any cause listed as kidney-related. This outcome was selected to align with the meta-analysis of the SGLT2 inhibitor trial individual participant data [1]. Secondary outcomes comprised a similar composite of a ≥40% decline in eGFR or ESKD, progression to uACR ≥30 mg/mmol, diabetic ketoacidosis, amputation or mycotic genital infection.

### Risk score

We estimated the 3-year absolute risk of kidney disease progression using the CKD-PC risk score for ≥50% decline in eGFR [10]. This risk score uses the same routine clinical features as the risk score for ≥40% decline in eGFR or ESKD (https://ckdpcrisk.org/gfrdecline40/) [10]: age, sex, eGFR, uACR, systolic BP, HbA_1c_, BMI, smoking status, use of antihypertensive medications, use of oral glucose-lowering treatments, use of insulin, previous diagnosis of atrial fibrillation, and previous diagnosis of heart failure or coronary heart disease (see electronic supplementary material [ESM] Methods) and is highly correlated (Pearson’s *r* 0.94) with that score [10].

### Covariates

The covariates included age (years), sex (self-reported; male/female), ethnicity (self-reported, categorised into major UK groups: White, South Asian, Black, Mixed, and other or unknown), IMD quintile, BMI (kg/m^2^), systolic BP (mmHg), total cholesterol (mmol/l), HbA_1c_ (mmol/mol and %), eGFR (calculated using the Chronic Kidney Disease Epidemiology Collaboration formula [21]; ml/min per 1.73 m^2^), uACR (mg/mmol), duration of type 2 diabetes (years), smoking status (non-smoker, ex-smoker or current smoker), previous diagnosis of arterial hypertension or atrial fibrillation, hospitalisation in the year before baseline, calendar year at baseline, number of current glucose-lowering treatments including insulin (1, 2 and ≥3), concurrent prescriptions for statins, insulin and ACE inhibitors or angiotensin II receptor blockers (full covariate set). For physiological and laboratory measurements, the value taken at the date closest to baseline in the previous 2 years was used. For subgroup analyses, low-level albuminuria (uACR 3–30 mg/mmol) was defined as having two consecutive uACR readings that met this threshold.

### Statistical analyses

This study comprised four main analytic steps. First, we conducted a real-world comparative effectiveness analysis to assess whether the relative treatment effect of SGLT2 inhibitors on kidney disease progression observed in a previous trial meta-analysis was applicable in the study population, including consistency across albuminuria levels (see ESM Table 1 for the target trial emulation protocol). Second, we evaluated the performance of the CKD-PC risk score for predicting kidney disease progression. Third, we integrated the relative treatment effect with the risk score to estimate individual-level absolute risk reductions for SGLT2 inhibitor treatment and assessed the accuracy of these predictions. Finally, we compared the clinical utility of a model-based treatment strategy to that of one based on a uACR threshold ≥3 mg/mmol. Secondary safety outcomes were also evaluated as part of the comparative effectiveness analysis.

**Table 1.**
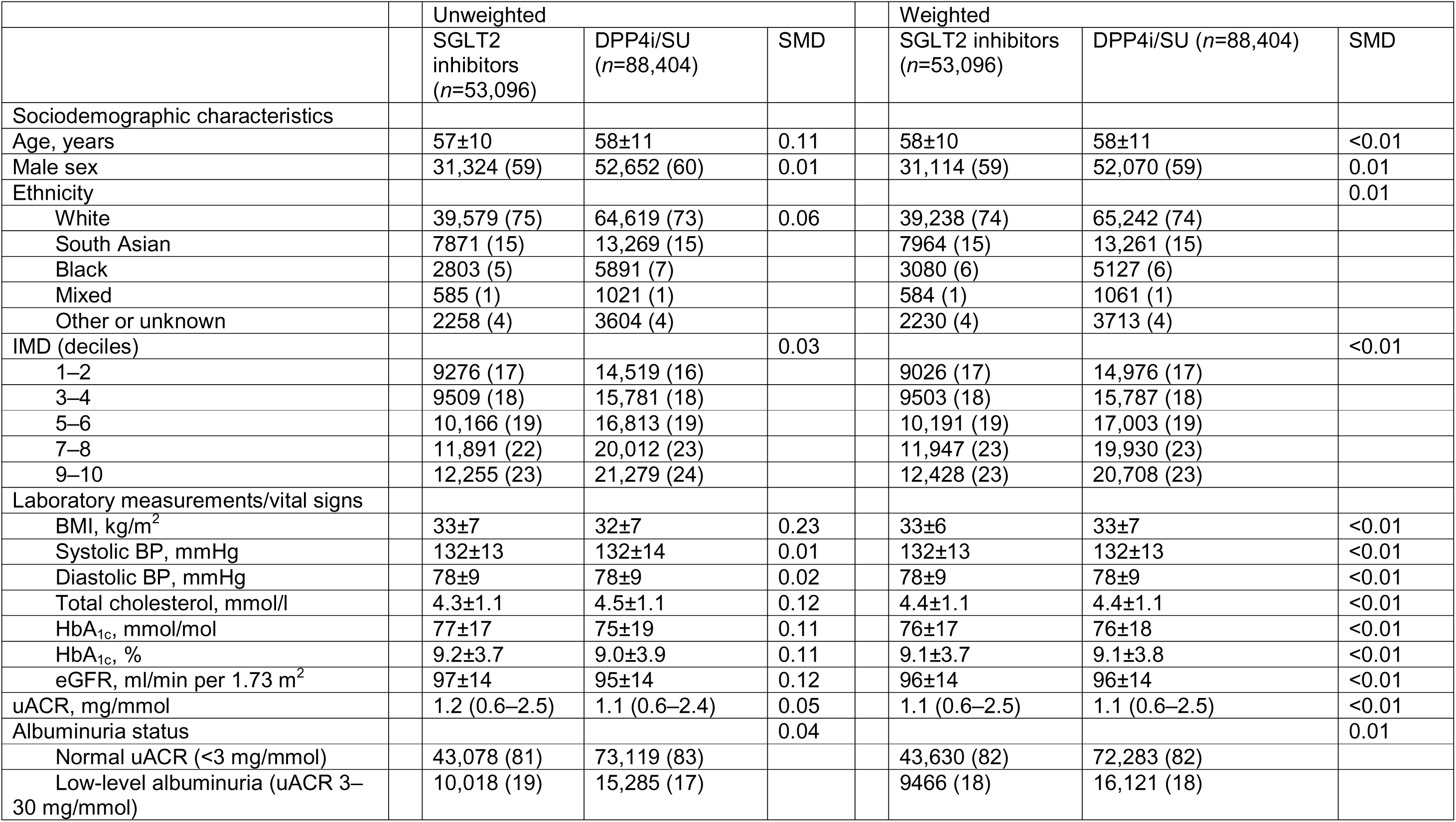
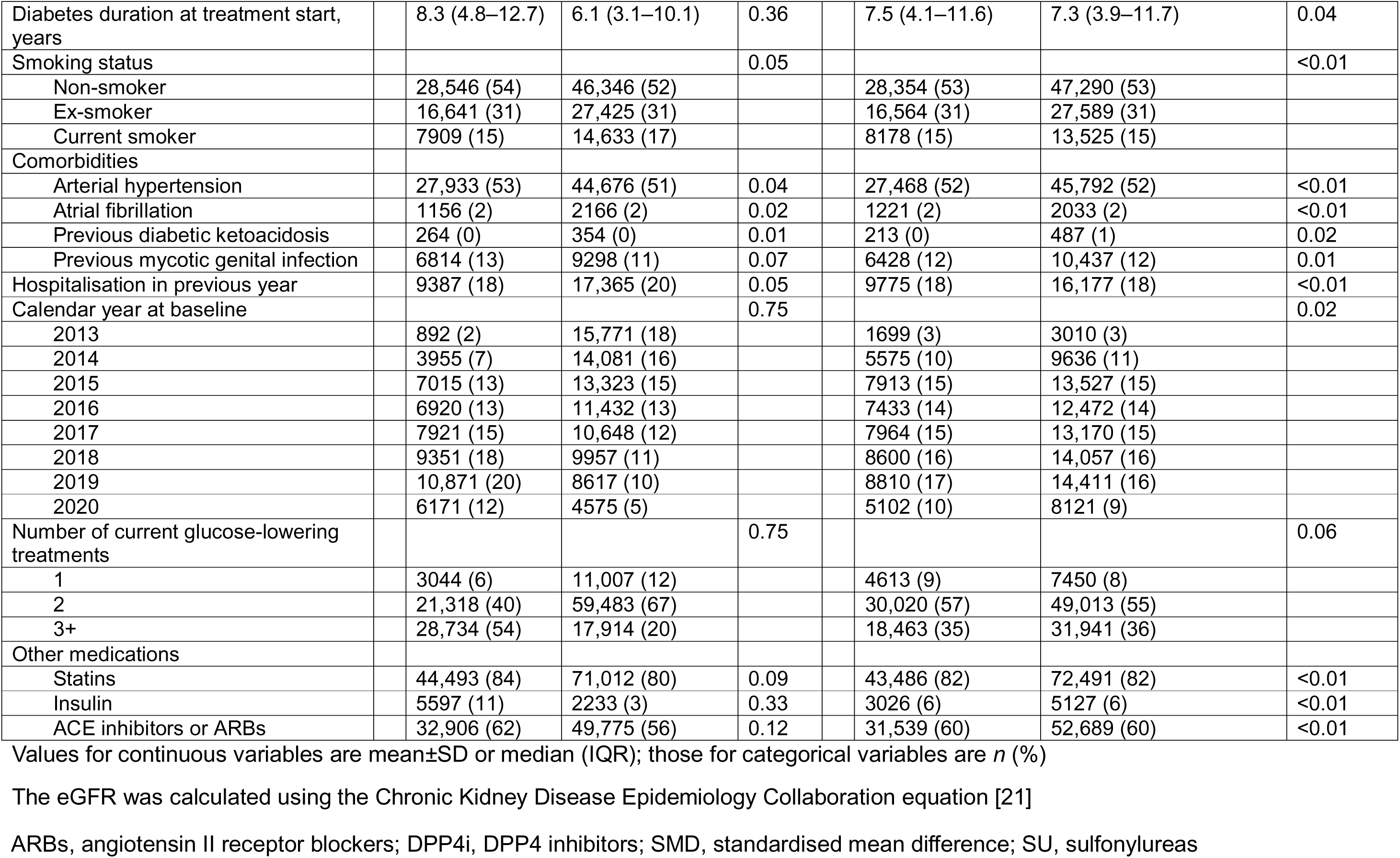
Baseline characteristics by treatment arm: unweighted and after overlap weighting.

|  | Unweighted |  |  | Weighted |  |  |
| --- | --- | --- | --- | --- | --- | --- |
|  | SGLT2 inhibitors<br>(*n*=53,096) | DPP4i/SU<br>(*n*=88,404) | SMD | SGLT2 inhibitors<br>(*n*=53,096) | DPP4i/SU (*n*=88,404) | SMD |
| Sociodemographic characteristics |  |  |  |  |  |  |
| Age, years | 57±10 | 58±11 | 0.11 | 58±10 | 58±11 | <0.01 |
| Male sex | 31,324 (59) | 52,652 (60) | 0.01 | 31,114 (59) | 52,070 (59) | 0.01 |
| Ethnicity |  |  |  |  |  | 0.01 |
| White | 39,579 (75) | 64,619 (73) | 0.06 | 39,238 (74) | 65,242 (74) |  |
| South Asian | 7871 (15) | 13,269 (15) |  | 7964 (15) | 13,261 (15) |  |
| Black | 2803 (5) | 5891 (7) |  | 3080 (6) | 5127 (6) |  |
| Mixed | 585 (1) | 1021 (1) |  | 584 (1) | 1061 (1) |  |
| Other or unknown | 2258 (4) | 3604 (4) |  | 2230 (4) | 3713 (4) |  |
| IMD (deciles) |  |  | 0.03 |  |  | <0.01 |
| 1–2 | 9276 (17) | 14,519 (16) |  | 9026 (17) | 14,976 (17) |  |
| 3–4 | 9509 (18) | 15,781 (18) |  | 9503 (18) | 15,787 (18) |  |
| 5–6 | 10,166 (19) | 16,813 (19) |  | 10,191 (19) | 17,003 (19) |  |
| 7–8 | 11,891 (22) | 20,012 (23) |  | 11,947 (23) | 19,930 (23) |  |
| 9–10 | 12,255 (23) | 21,279 (24) |  | 12,428 (23) | 20,708 (23) |  |
| Laboratory measurements/vital signs |  |  |  |  |  |  |
| BMI, kg/m<sup>2</sup> | 33±7 | 32±7 | 0.23 | 33±6 | 33±7 | <0.01 |
| Systolic BP, mmHg | 132±13 | 132±14 | 0.01 | 132±13 | 132±13 | <0.01 |
| Diastolic BP, mmHg | 78±9 | 78±9 | 0.02 | 78±9 | 78±9 | <0.01 |
| Total cholesterol, mmol/l | 4.3±1.1 | 4.5±1.1 | 0.12 | 4.4±1.1 | 4.4±1.1 | <0.01 |
| HbA<sub>1c</sub>, mmol/mol | 77±17 | 75±19 | 0.11 | 76±17 | 76±18 | <0.01 |
| HbA<sub>1c</sub>, % | 9.2±3.7 | 9.0±3.9 | 0.11 | 9.1±3.7 | 9.1±3.8 | <0.01 |
| eGFR, ml/min per 1.73 m<sup>2</sup> | 97±14 | 95±14 | 0.12 | 96±14 | 96±14 | <0.01 |
| uACR, mg/mmol | 1.2 (0.6–2.5) | 1.1 (0.6–2.4) | 0.05 | 1.1 (0.6–2.5) | 1.1 (0.6–2.5) | <0.01 |
| Albuminuria status |  |  | 0.04 |  |  | 0.01 |
| Normal uACR (<3 mg/mmol) | 43,078 (81) | 73,119 (83) |  | 43,630 (82) | 72,283 (82) |  |
| Low-level albuminuria (uACR 3–30 mg/mmol) | 10,018 (19) | 15,285 (17) |  | 9466 (18) | 16,121 (18) |  |
| Diabetes duration at treatment start, years | 8.3 (4.8–12.7) | 6.1 (3.1–10.1) | 0.36 | 7.5 (4.1–11.6) | 7.3 (3.9–11.7) | 0.04 |
| Smoking status |  |  | 0.05 |  |  | <0.01 |
| Non-smoker | 28,546 (54) | 46,346 (52) |  | 28,354 (53) | 47,290 (53) |  |
| Ex-smoker | 16,641 (31) | 27,425 (31) |  | 16,564 (31) | 27,589 (31) |  |
| Current smoker | 7909 (15) | 14,633 (17) |  | 8178 (15) | 13,525 (15) |  |
| Comorbidities |  |  |  |  |  |  |
| Arterial hypertension | 27,933 (53) | 44,676 (51) | 0.04 | 27,468 (52) | 45,792 (52) | <0.01 |
| Atrial fibrillation | 1156 (2) | 2166 (2) | 0.02 | 1221 (2) | 2033 (2) | <0.01 |
| Previous diabetic ketoacidosis | 264 (0) | 354 (0) | 0.01 | 213 (0) | 487 (1) | 0.02 |
| Previous mycotic genital infection | 6814 (13) | 9298 (11) | 0.07 | 6428 (12) | 10,437 (12) | 0.01 |
| Hospitalisation in previous year | 9387 (18) | 17,365 (20) | 0.05 | 9775 (18) | 16,177 (18) | <0.01 |
| Calendar year at baseline |  |  | 0.75 |  |  | 0.02 |
| 2013 | 892 (2) | 15,771 (18) |  | 1699 (3) | 3010 (3) |  |
| 2014 | 3955 (7) | 14,081 (16) |  | 5575 (10) | 9636 (11) |  |
| 2015 | 7015 (13) | 13,323 (15) |  | 7913 (15) | 13,527 (15) |  |
| 2016 | 6920 (13) | 11,432 (13) |  | 7433 (14) | 12,472 (14) |  |
| 2017 | 7921 (15) | 10,648 (12) |  | 7964 (15) | 13,170 (15) |  |
| 2018 | 9351 (18) | 9957 (11) |  | 8600 (16) | 14,057 (16) |  |
| 2019 | 10,871 (20) | 8617 (10) |  | 8810 (17) | 14,411 (16) |  |
| 2020 | 6171 (12) | 4575 (5) |  | 5102 (10) | 8121 (9) |  |
| Number of current glucose-lowering treatments |  |  | 0.75 |  |  | 0.06 |
| 1 | 3044 (6) | 11,007 (12) |  | 4613 (9) | 7450 (8) |  |
| 2 | 21,318 (40) | 59,483 (67) |  | 30,020 (57) | 49,013 (55) |  |
| 3+ | 28,734 (54) | 17,914 (20) |  | 18,463 (35) | 31,941 (36) |  |
| Other medications |  |  |  |  |  |  |
| Statins | 44,493 (84) | 71,012 (80) | 0.09 | 43,486 (82) | 72,491 (82) | <0.01 |
| Insulin | 5597 (11) | 2233 (3) | 0.33 | 3026 (6) | 5127 (6) | <0.01 |
| ACE inhibitors or ARBs | 32,906 (62) | 49,775 (56) | 0.12 | 31,539 (60) | 52,689 (60) | <0.01 |

Values for continuous variables are mean±SD or median (IQR); those for categorical variables are *n* (%)
The eGFR was calculated using the Chronic Kidney Disease Epidemiology Collaboration equation [21]
ARBs, angiotensin II receptor blockers; DPP4i, DPP4 inhibitors; SMD, standardised mean difference; SU, sulfonylureas

Missing clinical data (IMD 0.1%, BMI 3.1% [either height or weight missing], BP 0.2%, total cholesterol 0.3%, HbA_1c_ 0.2%, diabetes duration 5.8%, smoking status 0.6%) were imputed ten times using multivariate imputations with chained equations. IMD and smoking status were imputed using multinomial regression, BMI was imputed using passive imputation, and all other variables were imputed using predictive mean matching. We performed all analyses in each individual imputed dataset and pooled the results according to Rubin’s rules [22]. All analyses were performed using R version 4.4.1 (R Foundation for Statistical Computing, Vienna, Austria). We followed TRIPOD+AI guidance [23].

### Relative risk of kidney disease progression with SGLT2 inhibitor treatment

We estimated the relative risk of kidney disease progression when treated with SGLT2 inhibitors vs that for DPP4-inhibitors/sulfonylureas in our study population using doubly robust overlap-weighted Cox proportional hazards models[24, 25]. We tested for interaction by albuminuria status (uACR <3 vs 3–30 mg/mmol), and explored whether use of the continuous CKD-PC risk score modified the relative risk for SGLT2 inhibitors, with the risk score being fitted as a non-linear term (three-knot restricted cubic spline) interacting with treatment (adjusting for the full covariate set). As sensitivity analyses, we tested alternative adjustment methods, including doubly robust inverse probability of treatment weighted analyses (weights truncated at the 2nd and 98th percentiles) [26] and multivariable adjustment (full covariate set).

Weights were derived from a multivariable logistic regression-based propensity score model including the full covariate set. To check the validity of treating DPP4 inhibitors and sulfonylureas as a single comparator arm, we estimated the relative risk of kidney disease progression for each pairwise comparison of SGLT2 inhibitors, DPP4 inhibitors and sulfonylureas, using Cox proportional hazards models as described above. In these analyses, individuals were censored if they started treatment with any of the other study drugs.

### Performance of the CKD-PC risk score

We assessed calibration of the CKD-PC risk score [10] for predicting the primary outcome in our study population. We visually compared predicted and observed outcome proportions per risk score decile, and fitted a Cox proportional hazards model with the linear predictor as the only variable, estimating calibration-in-the-large (baseline hazard) and calibration slope [27]. Overall fit was assessed using the Brier score, and discrimination was assessed using the C statistic, with confidence intervals derived from 500 bootstrap samples.

### Derivation and validation of predicted 3-year absolute risk reductions

As calibration of the CKD-PC risk score was satisfactory (see Results), we used the risk score to estimate individual-level predicted absolute risk reductions from SGLT2 inhibitors (pARR) at 3 years as follows: *pARR* = S(0)t^HR^ - S(0)t, where HR represents the relative treatment effect (HR 0.62) from the SGLT2 inhibitor trial meta-analysis for individuals with diabetes overall [1], and S(0)t represents an individual’s predicted kidney disease progression-free survival at 3 years. Direct validation of *pARR* at the individual level was not possible, as counterfactual outcomes for each individual are unobservable, i.e. what would have happened had an individual received the alternative treatment. Building on our previous work validating models predicting differential glycaemic response to glucose-lowering medications [28], we assessed calibration of *pARR* using an approach tailored to time-to-event data. We estimated 3-year counterfactual absolute risk reductions for SGLT2 inhibitors based on the observed data using doubly robust overlap-weighted Cox proportional hazards models, incorporating treatment and covariates as previously described. We assessed calibration accuracy by visually comparing *pARR* to the counterfactual absolute risk reductions across deciles of pARR, and by estimating a calibration slope. As sensitivity analyses, we also assessed calibration of the model using only one comparator drug at a time (either DPP4 inhibitors or sulfonylureas).

### Comparison of treatment strategies based on albuminuria and pARR

We used decision curves to compare the clinical utility of two treatment strategies [29, 30]: a strategy based on uACR ≥3 mg/mmol (as recommended by current guidelines [2]) and a strategy based on pARR. For illustration, we chose a *pARR* threshold where a proportion of the population comparable to that for the albuminuria threshold would be treated. We modelled numbers treated, kidney disease progression events avoided and absolute risk reductions over 3 years by applying the risk score to individuals who would be treated and adjusting the risk score for those who would not be treated (incorporating the SGLT2 inhibitor relative treatment effect from the trial meta-analysis [1]). We further illustrated these comparisons in extended observational analyses with 5-year doubly robust overlap-weighted observed absolute risk reductions.

### Secondary outcomes

For secondary outcomes, we estimated relative risks with SGLT2 inhibitors vs DPP4 inhibitors/sulfonylureas when using the uACR threshold of ≥3 mg/mmol or *pARR* ≥0.65% using doubly robust overlap-weighted Cox proportional hazards models as described above.

### Ethics and data approval

Approval for the study was granted by the CPRD Independent Scientific Advisory Committee (eRAP 22_002000). The CPRD obtains annual research ethics approval from the UK’s Health Research Authority Research Ethics Committee (05/MRE04/87) to receive and supply patient data for public health research; no further ethical permissions were required for the analyses of these anonymised patient-level data.

### Patient and public involvement

Patients were not directly involved in the study design, but the project plan was presented to the Exeter Patient and Public Involvement Group, who advised on selection of a clinically relevant primary outcome.

## Results

### Study population

We included individuals with type 2 diabetes who received a first-ever prescription for SGLT2 inhibitors (*n*=53,096) or DPP4 inhibitors (*n*=53,521)/sulfonylureas (*n*=34,883) with preserved eGFR (≥60 ml/min per 1.73 m^2^) and normal uACR or low-level albuminuria (uACR 0–30 mg/mmol) and no prior history of heart failure or atherosclerotic CVD (ESM Fig. 1). Overall, the mean age was 58±11 years, and 83,976 of participants (59%) were men. Baseline characteristics by treatment arm are reported in Table 1. In the SGLT2 inhibitor arm, 10,018 (19%) had low-level albuminuria (uACR 3–30 mg/mmol), compared with 15,285 (17%) in the DPP4 inhibitor/sulfonylurea arm. Baseline characteristics by albuminuria status are reported in ESM Table 2.

**Fig. 1.**
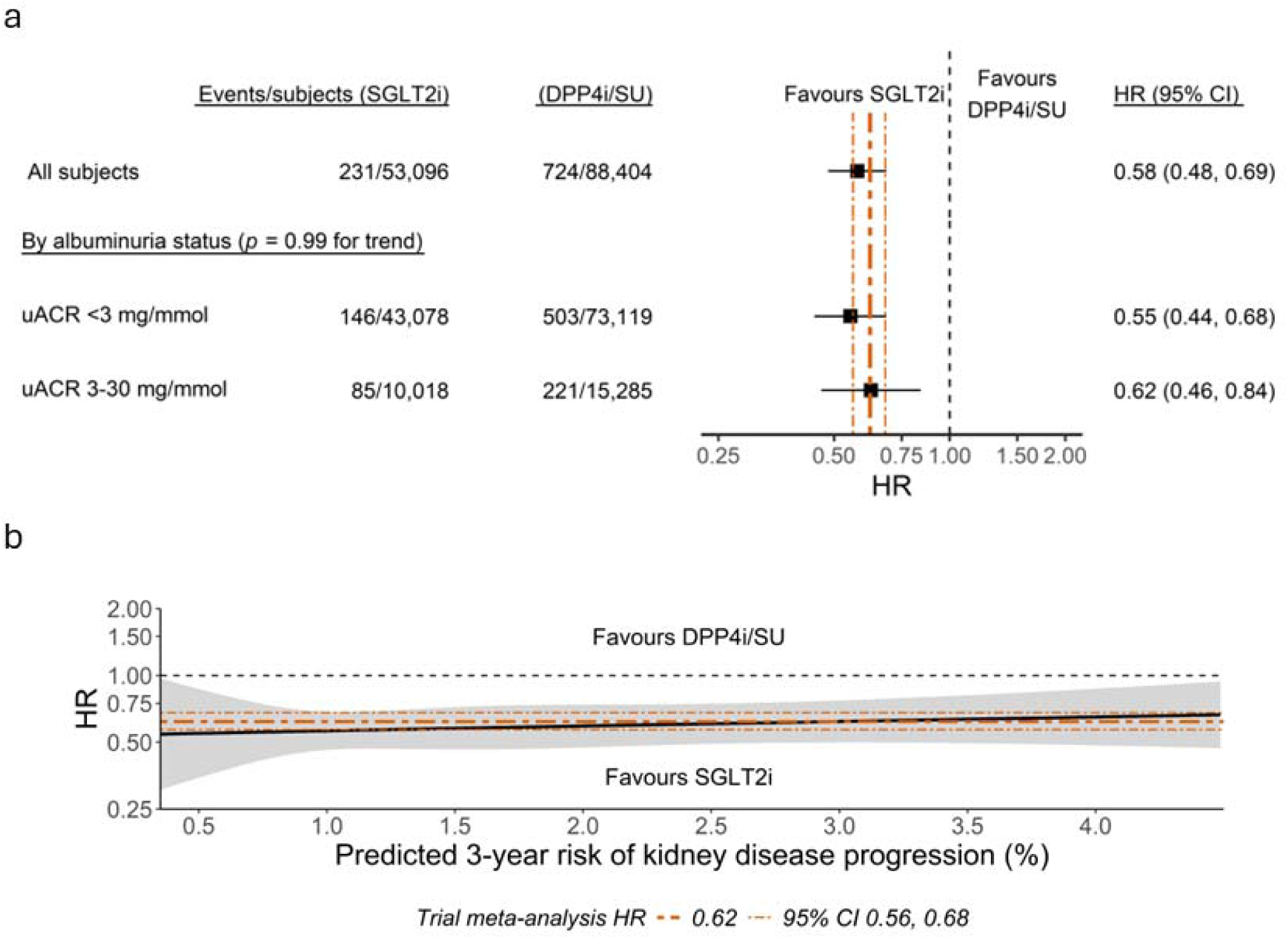
HR for kidney disease progression (≥50% decline in eGFR, ESKD or death due to kidney-related causes) in individuals receiving SGLT2 inhibitor treatment, by albuminuria status and CKD-PC risk score. (**a**) Forest plot with overlap-weighted HR for kidney disease progression over 3 years with SGLT2 inhibitors compared with DPP4i/SU (overall and by albuminuria status). (**b**) Continuous association between risk score (CKD-PC risk score for 3-year risk of kidney disease progression) and multivariable-adjusted HR for 3 year risk of kidney disease progression with SGLT2 inhibitors compared with DPP4i/SU, modelled using restricted cubic splines with three knots. For comparison, the trial meta-analysis HR and 95% CI are represented by orange dashed lines (HR 0.62; 95% CI 0.56, 0.68). DPP4i, DPP4 inhibitors; SGLT2i, SGLT2 inhibitors; SU, sulfonylureas

### Relative risk of kidney disease progression with SGLT2 inhibitor treatment

The incidence of kidney disease progression (≥50% decline in eGFR, ESKD, or death due to kidney-related causes) was lower with SGLT2 inhibitor treatment vs the comparator arm (2.2 vs 3.8 per 1000 person-years), with an overall 42% lower relative risk (overlap-weighted HR 0.58; 95% CI 0.48, 0.69). This reduction in relative risk was consistent for albuminuria status (*p*=0.99 for interaction, Fig. 1a) and continuous CKD-PC risk score (*p*=0.71 for non-linear and *p*=0.50 for linear risk score by treatment interaction term, Fig. 1b). Sensitivity analyses confirmed similar relative risks using inverse probability of treatment weighted or multivariable adjustment alone (ESM Fig. 2) and when comparing SGLT2 inhibitors to either DPP4 inhibitors or sulfonylureas separately (ESM Fig. 3). The lower relative risk with SGLT2 inhibitor treatment was similar to that in a previous SGLT2 inhibitor trial meta-analysis for the same outcome (HR 0.62; 95% CI 0.56, 0.68) [1], supporting the use of this estimate for predicting the real-world effect of SGLT2 inhibitor treatment in this population.

**Fig. 2.**
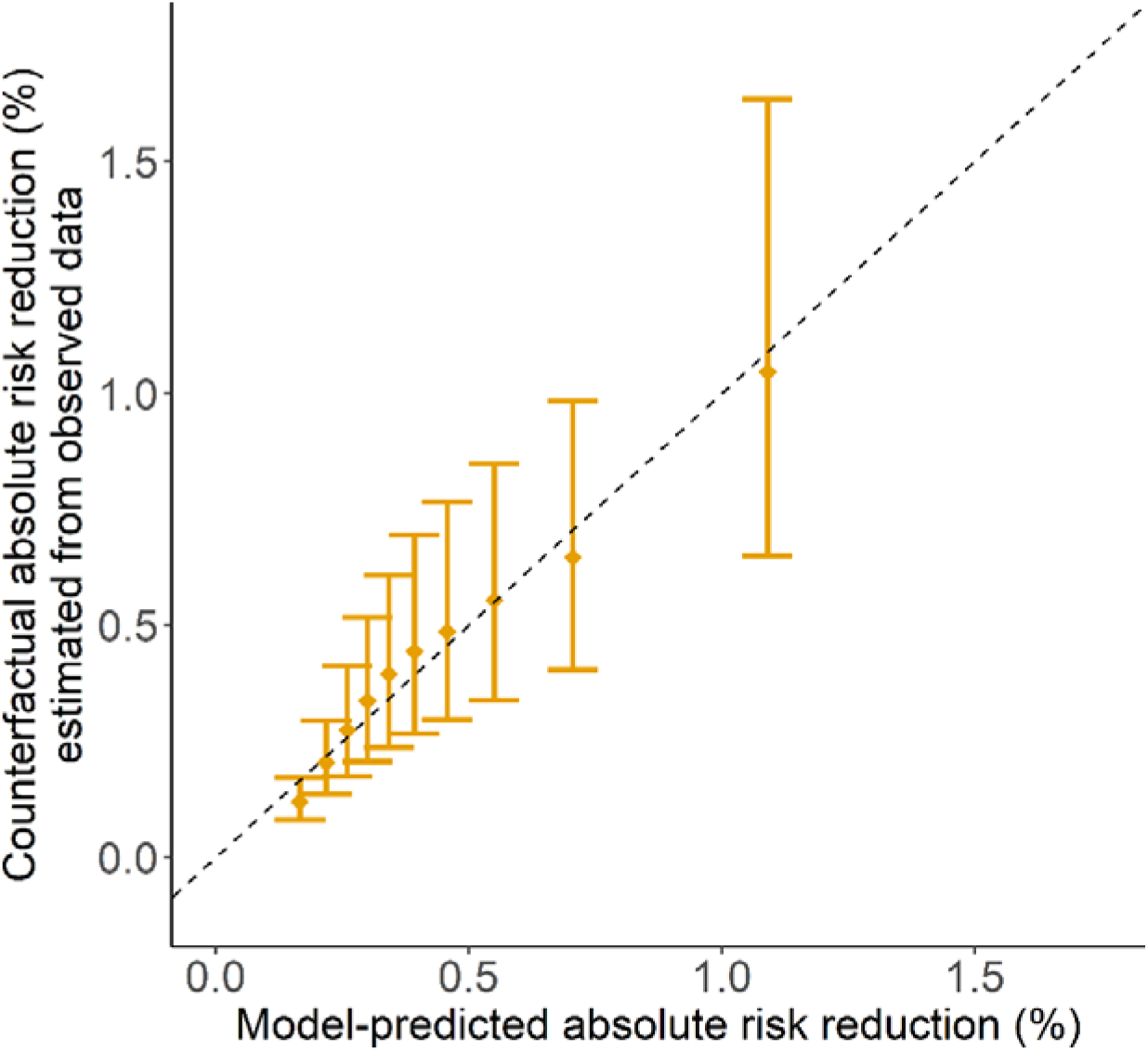
Calibration plot of model-predicted 3 year absolute risk reductions with SGLT2 inhibitor treatment (pARR) compared with counterfactual absolute risk reductions estimated from observed data using doubly robust overlap-weighted Cox models. Median and IQR are shown by deciles of pARR. Calibration plots with only one comparator drug at a time (either DPP4 inhibitors or sulfonylureas) are shown in ESM Fig. 5

**Fig. 3.**
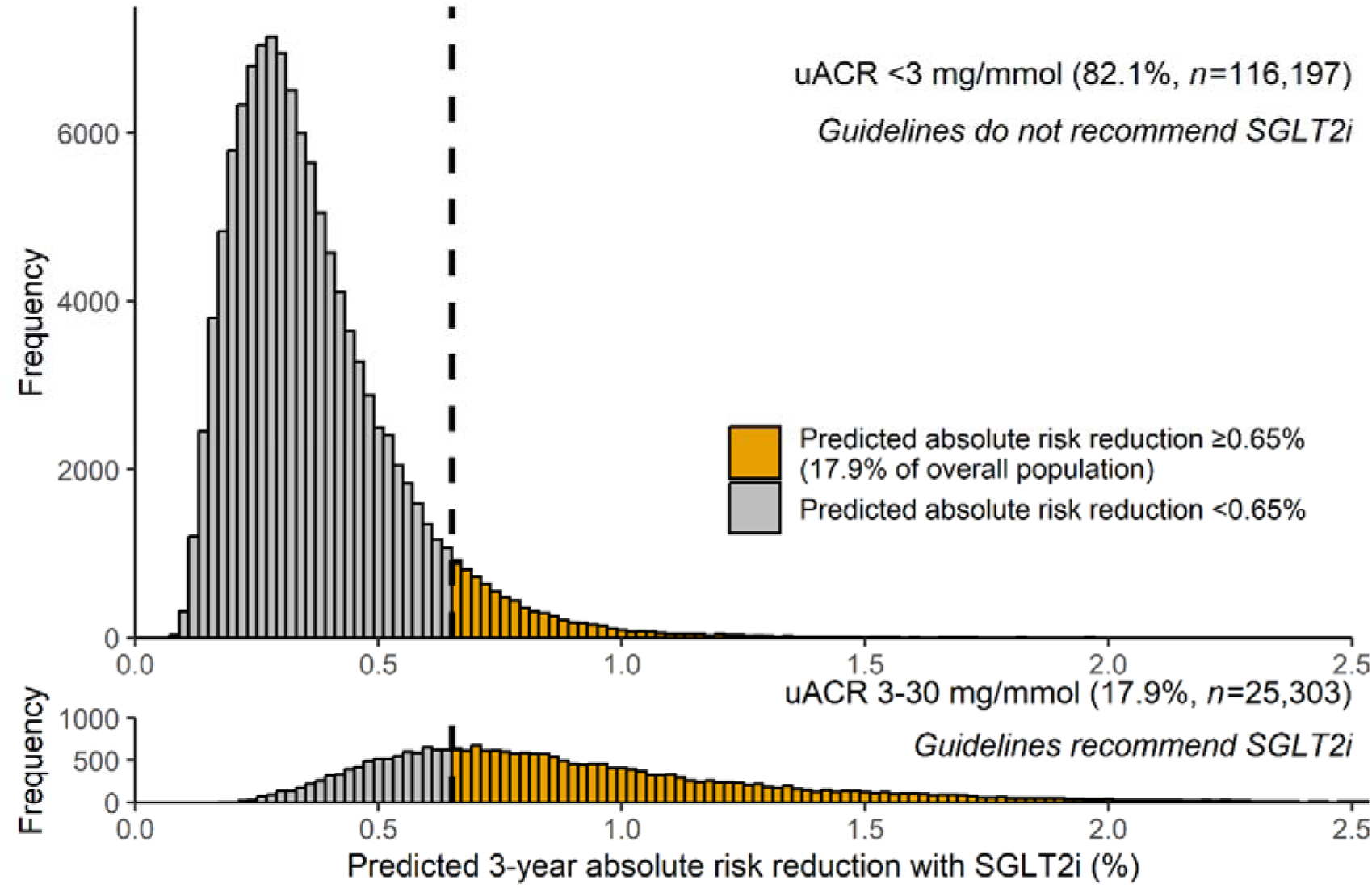
Distribution of predicted 3 year absolute risk reductions for SGLT2 inhibitor treatment (pARR) on kidney disease progression (≥50% decline in eGFR, ESKD or death due to kidney-related causes) in individuals with normal uACR (<3 mg/mmol; treatment not recommended by current guidelines) or low-level albuminuria (uACR 3–30 mg/mmol; treatment recommended by current guidelines). A vertical dashed line is shown at the ≥0.65% *pARR* threshold where a comparable proportion of the overall population would be targeted (17.9%) as under the ≥3 mg/mmol albuminuria threshold. SGLT2i, SGLT2 inhibitors

### Performance of the CKD-PC risk score

The overall prediction accuracy of the CKD-PC risk score for kidney disease progression was good (Brier score 0.0104; 95% CI 0.0097, 0.0111). The risk score showed good calibration (calibration-in-the-large 0.990; 95% CI 0.989, 0.991; calibration slope 1.05; 95% CI 0.94, 1.17; ESM Fig. 4), meaning that risk score recalibration was not required. Discrimination was modest (C statistic 0.68; 95% CI 0.67, 0.69). This was lower than the C statistic reported in the original validation study (which included people with severe albuminuria, heart failure and atherosclerotic CVD: C statistic 0.78; 95% CI 0.77, 0.79) but higher than that for uACR (continuous) in our population (C statistic 0.60; 95% CI 0.60, 0.61).

### Performance of combined *pARR* model

The predicted 3-year absolute risk reduction (pARR) for SGLT2 inhibitors, estimated by integrating the relative treatment effect from the SGLT2 inhibitor trial meta-analysis (HR 0.62) with the CKD-PC risk score, showed good calibration accuracy (Fig. 2), with a calibration slope of 1.10 (95% CI 1.09, 1.12). In the study population overall, the median *pARR* was 0.37% (IQR 0.26–0.55). A prototype decision support tool based on the *pARR* model is available at https://thijsjansz.shinyapps.io/shinyapp.

Figure 3 shows the distributions of *pARR* in individuals with normal uACR (<3 mg/mmol) and low-level albuminuria (uACR 3–30 mg/mmol). *pARR* was higher in the latter group (median 0.81%, IQR 0.60–1.10 vs median 0.33%, IQR 0.24–0.44). However, there was substantial variability in *pARR* within groups and overlap across both groups.

### Clinical utility of treatment strategies based on pARR

Decision curve analysis demonstrated that treatment strategies based on *pARR* consistently outperformed the ≥3 mg/mmol albuminuria threshold across all levels of risk tolerance (ESM Fig. 6). To illustrate this, we chose a *pARR* threshold of ≥0.65% to match the proportion of the population (17.9%) identified by the ≥3 mg/mmol albuminuria threshold (Fig. 3). Use of this *pARR* threshold resulted in an mean of 74 more true positives (treated individuals who developed kidney disease progression) and 75 more true negatives (non-treated individuals who did not develop kidney disease progression) per 100,000 individuals, compared with the ≥3 mg/mmol albuminuria threshold. In our study cohort, a treatment strategy based on *pARR* ≥0.65% would treat the same number of individuals as a strategy based on uACR ≥3 mg/mmol (*n*=25,303) but would prevent over 10% more events of kidney disease progression over 3 years (253 vs 228 events; ESM Table 3).

The treatment strategy based on *pARR* ≥0.65% identified a high-benefit subgroup within individuals with uACR <3 mg/mmol, comprising 6.7% of these individuals. These individuals were more likely to be female (56% vs 40%) and have higher systolic BP (mean 140 vs 131 mmHg), HbA_1c_ (mean 92 vs 73 mmol/mol or 10.6 vs 8.8%) and uACR (median 2.3 vs 0.8 mg/mmol; ESM Table 4). Extended observational analyses confirmed that among individuals with uACR <3 mg/mmol, those with above-threshold *pARR* also had a significantly higher benefit over 5 years compared to those with below-threshold *pARR* (observed overlap-weighted 5-year absolute risk reduction 3.2% vs 1.2%, *p*=0.05; ESM Figs 7 and 8). In contrast, among individuals with uACR 3–30 mg/mmol, the 30.7% with below-threshold *pARR* had a 5-year observed absolute risk reduction of only 1.1%, compared to 2.4% in those with above-threshold pARR.

### Secondary outcomes

The relative risks for ≥40% decline in eGFR/ESKD and progression to uACR ≥30 mg/mmol were consistently lower with SGLT2 inhibitors, whether stratified by albuminuria status or *pARR* (ESM Fig. 9). Relative risks of diabetic ketoacidosis and mycotic genital infection were higher with SGLT2 inhibitors (HR 1.92; 95% CI 1.51, 2.44 and HR 3.03; 95% CI 2.85, 3.03, respectively) and followed similar patterns when stratified by albuminuria status or pARR. There was no evidence of increased amputation risk.

## Discussion

Our model adapting the CKD-PC risk score accurately predicts the kidney protection benefit of SGLT2 inhibitors in individuals with type 2 diabetes, and no CKD or early-stage CKD (preserved eGFR and normal uACR or low-level albuminuria), who represent approximately 75% of those with type 2 diabetes. This approach leverages an internationally validated risk score and gold-standard evidence from a trial meta-analysis, providing a highly generalisable low-cost precision medicine tool. Use of a model-based treatment strategy could optimise resource utilisation and improve kidney outcomes: our analyses show that targeting the same number of individuals as the ≥3 mg/mmol albuminuria threshold currently recommended in international guidelines could prevent over 10% more kidney disease progression events, while identifying a previously unrecognised subgroup with normal uACR who could benefit substantially.

Our analysis suggests that the relative treatment effect of SGLT2 inhibitors from the trial meta-analysis [1] also applies to the lower-risk population that we evaluated. Prior analyses of randomised trials demonstrated consistent effects by eGFR and albuminuria status [1, 7–9], but did not evaluate whether this extends to individuals without heart failure or atherosclerotic CVD – a gap that our study addressed. We additionally found no evidence of heterogeneity in relative treatment effect across the spectrum of kidney disease progression risk. Furthermore, our analyses suggested consistent effects across secondary outcomes such as diabetic ketoacidosis and mycotic genital infection. Together, our results reinforce the findings of prior analyses showing that SGLT2 inhibitors consistently reduce the relative risk of kidney disease progression, irrespective of an individual’s CKD stage or progression risk.

This is the first study to evaluate the absolute kidney protection benefit of SGLT2 inhibitor treatment in a population with no CKD or only early-stage CKD –approaching a primary prevention context. This population with no CKD or only early-stage CKD was not studied in SGLT2 inhibitor kidney outcome trials [4–6]. Although this population was enrolled in cardiovascular outcome trials, these trials lacked analysis of absolute risk reductions by specific eGFR and albuminuria subgroups [7, 8]. Our study highlights a wide range of absolute risk reductions in this population, and presents a model that accurately predicts the individual-level kidney protection benefit of SGLT2 inhibitor treatment, which could enable individualised prescribing akin to that for statin treatment based on predicted cardiovascular benefit [12, 13].

Use of model-based treatment strategies could improve kidney outcomes and resource use by better aligning treatment with individual benefit. An example strategy that we evaluated could prevent 10% more kidney disease progression events than using the ≥3 mg/mmol albuminuria threshold, while identifying a hitherto unrecognised subgroup of individuals with normal uACR who could benefit substantially (predicted 3-year absolute risk reduction of 0.8%). For context, the 3-year absolute risk reduction was 3.3% in the meta-analysis of kidney outcome trials [1], which enrolled individuals at substantially higher risk. While the absolute number of events prevented with use of a model-based treatment strategy compared with the uACR threshold may appear modest over 3 years, this difference increased in extended 5-year analyses, suggesting greater longer-term benefit with a model-based treatment strategy. Importantly, decision curve analysis demonstrated that model-based treatment strategies consistently outperformed the ≥3 mg/mmol albuminuria threshold across all predicted benefit thresholds. Benefit thresholds could hence be tailored to reflect cost-effectiveness criteria, health system priorities or individual patient preferences. Thus, the model provides a flexible framework to support targeted prescribing, while also quantifying the potential benefit for an individual patient, facilitating shared decision-making.

### Strengths

Our study has several strengths. The model integrates gold-standard evidence from a trial meta-analysis [1] with an internationally validated clinical prediction model [10]. It was validated using robust epidemiological methods, including overlap weighting (incorporating calendar year to account for prescribing trends), in large-scale population-representative data. As these data are entirely independent of those used to develop both components of the prediction model, this provides strong external validation. The model is therefore highly generalisable and broadly applicable to clinical practice. Furthermore, the model only requires routine clinical features (such as age, sex, eGFR, albuminuria, systolic BP and HbA_1c_), supporting generalisation across patient subgroups, including both sexes, and facilitating straightforward implementation. This enables provision of low-cost precision medicine, which could be especially valuable in low-or middle-income countries, where type 2 diabetes prevalence is high but healthcare resources must be carefully prioritised [31]. While setting-specific optimisation may be required, the robust performance of the risk score suggests that, if required, only simple recalibration of baseline risks is likely to be necessary, rather than alterations to the model itself.

### Limitations

Our study has some limitations. First, the CKD-PC risk score showed more modest discrimination than in the original validation study [10], probably due to exclusion of individuals with severe albuminuria. Despite this, the *pARR* model showed greater discrimination and clinical utility than the currently recommended albuminuria threshold, highlighting the model’s added value in practice [32]. Second, we excluded individuals over 80 years old from the present analyses. Future research in that population should also account for competing risk of death. Third, follow-up for our primary analysis was limited to 3 years due to the risk score’s 3-year prediction window [10], and to align with most clinical trials [1], although our observational analyses showed that treatment differences were maintained for up to 5 years. Longer-term studies are needed to explore lifetime benefit. Finally, treatment allocation was non-random in this study, introducing potential for unmeasured confounding and indication bias.

### Future work

Future research should extend this work in several important areas. First, health economic analyses are needed to identify the most cost-effective *pARR* thresholds for treatment. Second, future studies should include individuals with eGFR 45–60 ml/min per 1.73 m^2^ and normal uACR or low-level albuminuria, who are recommended SGLT2 inhibitor treatment for kidney protection but who are not included in kidney outcome trials [2, 4–6]. We did not examine this group due to limited SGLT2 inhibitor use during the study period. Furthermore, the impact of acute eGFR changes when initiating treatment on long-term outcomes is unclear and should be evaluated in future work. Third, while our study focused on kidney outcomes, SGLT2 inhibitors also provide cardiovascular and glycaemic benefits [1]. Ultimately, future work should develop a comprehensive framework for predicting the individual-level benefit of all relevant outcomes. Fourth, emerging treatments such as glucagon-like peptide 1 receptor agonists have shown promise in reducing kidney disease progression risk [20], and future studies should assess their added benefit in individuals already being treated with SGLT2 inhibitors. Finally, integrating further predictors into our prediction model, such as genetic factors and novel biomarkers, could further improve precision, although at the expense of low-cost implementation.

### Conclusion

In conclusion, a risk score-based approach can predict the individual-level kidney protection benefit of SGLT2 inhibitor treatment. This enables individualised prescribing, similar to how predicted cardiovascular risk guides statin treatment for primary prevention, and could offer a more precise and effective alternative to the albuminuria threshold currently recommended in international guidelines. As the prediction model only uses routine clinical features, it is straightforward to implement in clinical practice, providing a low-cost, globally applicable precision medicine tool for people with type 2 diabetes.

## Data Availability

CPRD data are available by application to the CPRD Independent Scientific Advisory Committee. All R code to reproduce the analyses in this paper is available at https://github.com/Exeter-Diabetes/CPRD-Thijs-SGLT2-KF-scripts

https://github.com/Exeter-Diabetes/CPRD-Codelists

https://github.com/Exeter-Diabetes/CPRD-Cohort-scripts

## Abbreviations

CKD: Chronic kidney disease
CKD-PC: Chronic Kidney Disease Prognosis Consortium
DPP4: Dipeptidyl peptidase-4
ESKD: End-stage kidney disease
IMD: Index of Multiple Deprivation
SGLT2: Sodium–glucose cotransporter-2
uACR: Urinary albumin/creatinine ratio

## Acknowledgements

This article is based in part on data from the Clinical Practice Research Datalink (CPRD) Aurum obtained under licence from the UK Medicines and Healthcare Products Regulatory Agency. CPRD data are provided and collected by the UK National Health Service as part of patient care and support. This research was carried out at the National Institute for Health and Care Research (NIHR) Exeter Biomedical Research Centre (BRC). The views expressed are those of the authors and not necessarily those of the NIHR or the Department of Health and Social Care.

## Data availability

All CPRD data are available by application to the CPRD Independent Scientific Advisory Committee (https://cprd.com/data-access).

## Code availability

All code to develop the CPRD cohort used in this study is available at https://github.com/Exeter-Diabetes/CPRD-Codelists/tree/pre-2024. Code to reproduce the analyses in this article is available at https://github.com/Exeter-Diabetes/CPRD-Thijs-SGLT2-KF-scripts.

## Funding

This research was supported by the Medical Research Council (UK) (MR/N00633X/1) and the European Foundation for the Study of Diabetes. The funders had no role in the study design, data collection, data analysis, data interpretation, writing of the article, or the decision to submit for publication.

## Authors’ relationships and activities

JMD is supported by a Wellcome Trust Early Career award (227070/Z/23/Z). ATH and BMS are supported by the NIHR Exeter Clinical Research Facility; the views expressed are those of the authors and not necessarily those of the NHS, the NIHR or the Department of Health. AGJ declares research funding to his university from the UK Medical Research Council, NIHR, Diabetes UK, Breakthrough T1D, the Novo Nordisk Foundation and European Foundation for the Study of Diabetes. ERP has received honoraria for speaking from Lilly, Novo Nordisk and Illumina. RAO declares consulting income from Sanofi and honoraria from Sanofi and Novo Nordisk, in addition to research funding from NIDDK, Breakthrough T1D, The Leona M. and Larry B. Helmsley Charitable Trust and Randox Ltd. No industry representatives were involved in the writing of the manuscript or analysis of data. For all authors these relationships are outside the submitted work; there are no other relationships or activities that might bias, or be perceived to bias, their work.

## Contribution statement

The study concept and design were conceived and developed by TTJ, KGY, BMS, ATH, AGJ, RAO and JMD. TTJ, KGY, RH, BMS and JMD had access to all the raw datasets used for the study. TTJ directly accessed the Clinical Practice Research Datalink (CPRD), with support from KGY, RH, BMS and JMD. TTJ, KGY and JMD accessed and verified the underlying data used in the study. TTJ developed the kidney protection benefit model online calculator. All authors had full access to the complete results of the analysis of the CPRD dataset. All authors provided support for the analysis and interpretation of results, critically revised the article, and read and approved the final article. TTJ and JMD attest that all listed authors meet authorship criteria and that no others meeting the criteria have been omitted. TTJ is responsible for the integrity of the work as a whole. All authors had final responsibility for the decision to submit for publication.

## Notes

### Funding Statement

This research was supported the Medical Research Council (UK) (MR/N00633X/1). The funders had no role in the study design, data collection, data analysis, data interpretation, writing of the article, or the decision to submit for publication.

### Author Declarations

Approval for the study was granted by the CPRD Independent Scientific Advisory Committee (eRAP 22_002000) and Vivli (ID 00005959). CPRD obtains annual research ethics approval from the UK's Health Research Authority Research Ethics Committee (05/MRE04/87) to receive and supply patient data for public health research; no further ethical permissions were required for the analyses of these anonymised patient-level data.

### Summary of Updates

Final version of manuscript as accepted by Diabetologia.

