## Supplemental Material for "Precision medicine in type 2 diabetes: targeting SGLT2 inhibitor treatment for kidney protection"

**ELECTRONIC SUPPLEMENTARY MATERIAL (ESM)**


| Item | Page |
|---|---|
| ESM Methods | 2 |
| ESM Table 1 | 3 |
| ESM Table 2 | 6 |
| ESM Table 3 | 9 |
| ESM Table 4 | 10 |
| ESM Figure 1 | 13 |
| ESM Figure 2 | 14 |
| ESM Figure 3 | 15 |
| ESM Figure 4 | 16 |
| ESM Figure 5 | 17 |
| ESM Figure 6 | 19 |
| ESM Figure 7 | 20 |
| ESM Figure 8 | 21 |
| ESM Figure 9 | 22 |

**ESM Methods**

The formula for the CKD-PC risk score formula for ≥50% eGFR decline/ESKD is as follows:

$$\frac{e^{(-4.941086+linea\quad predictor)}}{1+e^{(-4.941086+line\quad predictor)}}$$

Where the linear predictor is defined as follows:

$$\begin{aligned}
&0.0794321 \times ((age\ [in\ years] - 60)/10) - 0.1141482 \times (presence\ of\ male\ sex\ [binary] - 0.5) \\
&- 0.0459708 \times ((eGFR\ [in\ mL/min/1.73m2] - 85)/5 + 0.4326073 \times \ln(uACR\ [in\ mg/g]\ /\ 10) \\
&+ 0.1917602 \times ((systolic\ blood\ pressure\ [in\ mmHg] - 130)\ /\ 20) \\
&+ 0.306044 \times use\ of\ antihypertensive\ medication\ [binary] - 0.0670274 \times ((systolic\ blood\ pressure\ [in\ mmHg] \\
&- 130)\ /\ 20 \times use\ of\ antihypertensive\ medication\ [binary]) + 0.9185919 \times (history\ of\ heart\ failure\ [binary] - 0.05) \\
&+ 0.2386131 \times (history\ of\ coronary\ artery\ disease\ [binary] - 0.15) + 0.2157927 \times bistory\ of\ atrial\ fibrillation\ [binary] \\
&+ 0.1122236 \times current\ smoker\ [binary] + 0.1327815 \times former\ smoker\ [binary] + 0.0154271 \times ((BMI\ [in\ kg/m2] \\
&- 30)\ /\ 5) + 0.1103419 \times (HbA1c\ [in\ \%] - 7) - 0.0460205 \times use\ of\ oral\ antihyperglycaemic\ medication\ [binary] \\
&+ 0.2571656 \times use\ of\ insulin\ [binary]
\end{aligned}$$

Of note, history of heart failure (0), history of heart failure (0) and use of oral antihyperglycaemic medication (1) were constant in this study population.

**ESM Table 1.** Target trial emulation protocol.

| Protocol components | Target trial | Emulation |
|---|---|---|
| **Eligibility criteria** | **Inclusion**:<br>- Type 2 diabetes<br>- SGLT2-inhibitor naïve<br><br>**Exclusion**:<br>- eGFR <60 ml/min/1.73m$^2$ or need for renal replacement therapy (dialysis or kidney transplant).<br>- Urinary albumin creatinine ratio ≥30 mg/mmol.<br>- Prior history of atherosclerotic vascular disease (i.e. myocardial infarction, stroke, or peripheral vascular disease) or heart failure.<br>- Use of GLP1-receptor agonists. | **Inclusion**<br>- Type 2 diabetes<br>- SGLT2-inhibitor naïve<br>- Receiving first-ever prescription for an SGLT2-inhibitor, DPP4-inhibitor or sulfonylurea.<br><br>**Exclusion**:<br>- eGFR <60 ml/min/1.73m$^2$ or need for renal replacement therapy (dialysis or kidney transplant).<br>- Urinary albumin creatinine ratio ≥30 mg/mmol.<br>- Prior history of atherosclerotic vascular disease (i.e. myocardial infarction, stroke, or peripheral vascular disease) or heart failure.<br>- Use of GLP1-receptor agonists.<br>- Less than 91 days of registration data before treatment initiation. |
| **Treatment assignment** | Random assignment to one of the two treatment arms (SGLT2-inhibitors vs DPP4-inhibitors/sulfonylureas). | We assume random assignment to one of the two treatment arms (SGLT2-inhibitors vs DPP4-inhibitors/sulfonylureas) by applying overlap weighting based on predefined covariates. |
| **Treatment initiation** | Initiation of only one antihyperglycaemic medication (SGLT2-inhibitor, DPP4-inhibitor, or sulfonylurea) at the time of treatment assignment. | We consider the date of a first prescription for one antihyperglycaemic medication (SGLT2-inhibitor, DPP4-inhibitor, or sulfonylurea) as date of treatment initiation. |

| | | |
|---|---|---|
| **Treatment strategy** | Prescription issued by primary care practice. Further additional treatment after a 3-month treatment initiation period is allowed as needed. | Prescription information identified from primary care records (Clinical Practice Research Datalink, CPRD). Further additional treatment after a 3-month treatment initiation period is allowed as needed. |
| **Follow-up period** | Follow-up starts at treatment initiation. Subjects are followed up until the earliest of: occurrence of outcome, a maximum of 3 years, or September 30, 2023. | Follow-up starts at treatment initiation. Subjects are followed up until the earliest of: occurrence of outcome, a maximum of 3 years, or September 30, 2023. Subjects in the DPP4-inhibitor/sulfonylurea arm are censored if starting an SGLT2-inhibitor or GLP1-receptor agonist. |
| **Outcomes** | Primary outcome:<br>- Kidney disease progression defined as a composite of a ≥50% decline in eGFR, end-stage kidney disease (requirement for renal replacement therapy or sustained eGFR <15 ml/min/1.73m$^2$), or death with kidney disease listed as cause of death.<br>Secondary outcomes:<br>- A similar composite of a ≥40% decline in eGFR or end-stage kidney disease<br>- Progression to uACR ≥30 mg/mmol<br>- Diabetic ketoacidosis<br>- Amputation<br>- Mycotic genital infection | Primary outcome:<br>- Kidney disease progression defined as a composite of a ≥50% decline in eGFR, end-stage kidney disease (requirement for renal replacement therapy or sustained eGFR <15 ml/min/1.73m$^2$), or death with kidney disease listed as cause of death.<br>Secondary outcomes:<br>- A similar composite of a ≥40% decline in eGFR or end-stage kidney disease<br>- Progression to uACR ≥30 mg/mmol<br>- Diabetic ketoacidosis<br>- Amputation<br>- Mycotic genital infection |
| **Causal contrasts of interest** | Average treatment effects in intention-to-treat analyses. | Average treatment effects in intention-to-treat analyses. |

| | | |
|---|---|---|
| **Analysis plan** | - Cox proportional hazard models.<br>- Subgroup analyses with stratification by presence of low-level albuminuria (≥3 mg/mmol). | - Doubly-robust Cox proportional hazards models (adjusted for predefined covariates and weighted for overlap weights).<br>- Subgroup analyses with stratification by presence of low-level albuminuria (≥3 mg/mmol).<br>- Sensitivity analyses with weighting for inverse probability of treatment weights instead of overlap weights, and multivariable adjustment alone.<br>- Sensitivity analyses with DPP4-inhibitors and sulfonylureas treated as separate treatment arms, with subjects censored if starting any of the other study drugs, analysed as described above. |

Abbreviations: SGLT2-inhibitors sodium-glucose cotransporter-2 inhibitors, DPP4-inhibitors dipeptidyl peptidase-4 inhibitors, GLP1-receptor agonists glucagon-like peptide-1 receptor agonists, eGFR estimated glomerular filtration rate.

**ESM Table 2.** Baseline characteristics by albuminuria status.

| | uACR <3 mg/mmol (*n*=116,197) | uACR ≥3 mg/mmol (*n*=25,303) |
|---|---|---|
| *Sociodemographic characteristics* | | |
| Age, years | 58 ±11 | 57 ±11 |
| Male sex | 67,999 (59%) | 15,977 (63%) |
| Ethnicity | | |
| White | 86,720 (75%) | 17,478 (69%) |
| South Asian | 16,435 (14%) | 4,705 (19%) |
| Black | 7047 (6%) | 1647 (7%) |
| Mixed | 1287 (1%) | 319 (1%) |
| Other or unknown | 4708 (4%) | 1154 (5%) |
| Index of multiple deprivation, deciles | | |
| 1-2 | 20,045 (17%) | 3750 (15%) |
| 3-4 | 21,047 (18%) | 4241 (17%) |
| 5-6 | 22,224 (19%) | 4756 (19%) |
| 7-8 | 25,919 (22%) | 5983 (24%) |
| 9-10 | 26,961 (23%) | 6573 (26%) |
| *Laboratory and vital signs measurements* | | |
| BMI, $kg/m^2$ | 32 ±7 | 33 ±7 |
| Systolic blood pressure, mmHg | 132 ±13 | 135 ±14 |
| Diastolic blood pressure, mmHg | 78 ±9 | 79 ±9 |
| Total cholesterol, mmol/l | 4.4 ±1.1 | 4.5 ±1.2 |
| $HbA_{1c}$, mmol/mol | 75 ±18 | 79 ±19 |
| $HbA_{1c}$, % | 9.0 ±3.8 | 9.4 ±3.9 |
| eGFR, $ml/min/1.73m^2$ | 95 ±14 | 97 ±15 |
| uACR, mg/mmol | 0.9 [0.6, 1.6] | 6.1 [4.1, 10.5] |
| *Comorbidities* | | |
| Diabetes duration at treatment start, years | 6.8 [3.6, 11.0] | 7.3 [3.8, 11.7] |

| | | |
|---|---|---|
| Smoking status | | |
| Non-smoker | 62,311 (54%) | 12,581 (50%) |
| Ex-smoker | 36,257 (31%) | 7816 (31%) |
| Current smoker | 17,630 (15%) | 4906 (19%) |
| Arterial hypertension | 57,873 (50%) | 14,736 (58%) |
| Atrial fibrillation | 2620 (2%) | 702 (3%) |
| Previous diabetic keto-acidosis | 465 (0%) | 153 (1%) |
| Previous mycotic genital infection | 13,239 (11%) | 2873 (11%) |
| Hospitalisation in previous year | 21,563 (19%) | 5189 (21%) |
| *Medications* | | |
| Calendar year at baseline | | |
| 2013 | 13,826 (12%) | 2837 (11%) |
| 2014 | 14,795 (13%) | 3241 (13%) |
| 2015 | 16,762 (14%) | 3576 (14%) |
| 2016 | 15,121 (13%) | 3231 (13%) |
| 2017 | 15,210 (13%) | 3359 (13%) |
| 2018 | 15,813 (14%) | 3495 (14%) |
| 2019 | 15,901 (14%) | 3587 (14%) |
| 2020 | 8769 (8%) | 1977 (8%) |
| Number of current glucose-lowering treatments | | |
| 1 | 11,897 (10%) | 2154 (9%) |
| 2 | 67,175 (58%) | 13,626 (54%) |
| 3+ | 37,125 (32%) | 9523 (38%) |
| On statin | 94,551 (81%) | 20,954 (83%) |
| On insulin | 5875 (5%) | 1955 (8%) |
| On ACE-inhibitor or ARB | 64,014 (55%) | 18,667 (74%) |
| Treatment arm | | |
| SGLT2-inhibitor | 43,078 (37%) | 10,018 (40%) |

| | | |
|---|---|---|
| DPP4-inhibitor/sulfonylurea | 73,119 (63%) | 15,285 (60%) |

Values for continuous variables are mean±SD or median (IQR); those for categorical variables are *n* (%)

Abbreviations: BMI body mass index, eGFR estimated glomerular filtration rate (calculated using the Chronic Kidney Disease Epidemiology Collaboration equation), uACR urinary albumin-creatinine ratio, ACE angiotensin-converting enzyme, ARB angiotensin-II receptor blocker, SGLT2-inhibitors sodium-glucose cotransporter-2 inhibitors, DPP4-inhibitors dipeptidyl peptidase-4 inhibitors.

**ESM Table 3.** Estimated number of kidney disease progression events over 3 years with different SGLT2-inhibitor treatment strategies modelled on the study population. Event numbers were estimated using the CKD-PC risk score and the relative treatment effect of SGLT2-inhibitors from previous trial meta-analysis (HR 0.62[1]).

| **Strategy** | ***N* (%) treated with SGLT2-inhibitor** | ***N* (%) with kidney disease progression event over 3 years** | ***N* events avoided (% of total avoidable)** | **3-year absolute risk reduction** |
|---|---|---|---|---|
| Treat none | 0 (0%) | 1,727 (1.2%) | 0 (0%) | - |
| Treat all | 141,500 (100%) | 1,075 (0.8%) | 652 (100%) | 0.46%<br>*(NNT 217)* |
| Albuminuria threshold ≥3 mg/mmol | 25,303 (17.9%) | 1,499 (1.1%) | 228 (35.0%) | 0.90%<br>*(NNT 111)* |
| $pARR$ threshold ≥0.65% | 25,303 (17.9%) | 1,474 (1.0%) | 253 (38.8%) | 1.00%<br>*(NNT 100)* |

Abbreviations: $pARR$ predicted 3-year absolute risk reduction with SGLT2-inhibitors, NNT number needed to treat.

**ESM Table 4.** Baseline characteristics by $pARR$ threshold.

| | uACR <3 mg/mmol | | uACR 3-30 mg/mmol | |
|---|---|---|---|---|
| | $pARR$ <0.65% (*n*=108,429) | $pARR$ ≥0.65% (*n*=7768) | $pARR$ <0.65% (*n*=7768) | $pARR$ ≥0.65% (*n*=17,535) |
| *Sociodemographic characteristics* | | | | |
| Age, years | 58 ±11 | 65 ±9 | 52 ±11 | 60 ±10 |
| Male sex | 64,584 (60%) | 3415 (44%) | 5059 (65%) | 10,918 (62%) |
| Ethnicity | | | | |
| White | 80,415 (74%) | 6305 (81%) | 4796 (62%) | 12,682 (72%) |
| South Asian | 15,836 (15%) | 599 (8%) | 1973 (25%) | 2732 (16%) |
| Black | 6491 (6%) | 556 (7%) | 453 (6%) | 1194 (7%) |
| Mixed | 1201 (1%) | 86 (1%) | 111 (1%) | 208 (1%) |
| Other or unknown | 4487 (4%) | 221 (3%) | 435 (6%) | 719 (4%) |
| Index of multiple deprivation, deciles | | | | |
| 1-2 | 18,729 (17%) | 1316 (17%) | 1100 (14%) | 2650 (15%) |
| 3-4 | 19,635 (18%) | 1412 (18%) | 1252 (16%) | 2989 (17%) |
| 5-6 | 20,730 (19%) | 1494 (19%) | 1410 (18%) | 3347 (19%) |
| 7-8 | 24,171 (22%) | 1749 (23%) | 1859 (24%) | 4125 (24%) |
| 9-10 | 25,165 (23%) | 1796 (23%) | 2148 (28%) | 4425 (25%) |
| *Laboratory and vital signs measurements* | | | | |
| BMI, kg/m$^2$ | 32 ±7 | 33 ±7 | 32 ±7 | 33 ±7 |
| Systolic blood pressure, mmHg | 131 ±13 | 140 ±16 | 129 ±12 | 137 ±15 |
| Diastolic blood pressure, mmHg | 78 ±8 | 79 ±10 | 79 ±8 | 79 ±10 |
| Total cholesterol, mmol/l | 4.4 ±1.1 | 4.6 ±1.2 | 4.5 ±1.2 | 4.5 ±1.2 |
| $HbA_{1c}$, mmol/mol | 73 ±16 | 92 ±23 | 72 ±15 | 82 ±20 |
| $HbA_{1c}$, % | 8.8 ±3.6 | 10.6 ±4.3 | 8.7 ±3.5 | 9.7 ±4.0 |
| eGFR, ml/min/1.73m$^2$ | 96 ±14 | 85 ±14 | 105 ±12 | 93 ±15 |
| uACR, mg/mmol | 0.8 [0.6, 1.4] | 2.3 [1.7, 2.8] | 4.1 [3.4, 5.4] | 7.8 [4.9, 13.0] |

| | | | | |
|---|---|---|---|---|
| *Comorbidities* | | | | |
| Diabetes duration at treatment start, years | 6.7 [3.6, 10.8] | 8.6 [4.6, 13.3] | 5.7 [2.9, 9.5] | 8.1 [4.4, 12.6] |
| Smoking status | | | | |
| Non-smoker | 59,143 (55%) | 3167 (41%) | 4588 (59%) | 7993 (46%) |
| Ex-smoker | 32,803 (30%) | 3453 (45%) | 1686 (22%) | 6130 (35%) |
| Current smoker | 16,483 (15%) | 1147 (15%) | 1495 (19%) | 3412 (20%) |
| Arterial hypertension | 51,941 (48%) | 5932 (76%) | 2925 (38%) | 11,811 (67%) |
| Atrial fibrillation | 1976 (2%) | 644 (8%) | 28 (0%) | 674 (4%) |
| Previous diabetic keto-acidosis | 410 (0%) | 55 (1%) | 34 (0%) | 119 (1%) |
| Previous mycotic genital infection | 12,200 (11%) | 1039 (13%) | 857 (11%) | 2016 (12%) |
| Hospitalisation in previous year | 19,868 (18%) | 1695 (22%) | 1513 (20%) | 3676 (21%) |
| *Medications* | | | | |
| Calendar year at baseline | | | | |
| 2013 | 12,923 (12%) | 903 (12%) | 911 (12%) | 1926 (11%) |
| 2014 | 13,893 (13%) | 902 (12%) | 958 (12%) | 2283 (13%) |
| 2015 | 15,607 (14%) | 1155 (15%) | 1068 (14%) | 2508 (14%) |
| 2016 | 14,038 (13%) | 1083 (14%) | 998 (13%) | 2233 (13%) |
| 2017 | 14,163 (13%) | 1047 (14%) | 1035 (13%) | 2324 (13%) |
| 2018 | 14,759 (14%) | 1054 (14%) | 1048 (14%) | 2447 (14%) |
| 2019 | 14,906 (14%) | 995 (13%) | 1166 (15%) | 2421 (14%) |
| 2020 | 8140 (8%) | 629 (8%) | 584 (8%) | 1393 (8%) |
| Number of current glucose-lowering treatments | | | | |
| 1 | 11,086 (10%) | 811 (10%) | 672 (9%) | 1482 (9%) |
| 2 | 63,400 (58%) | 3775 (49%) | 4767 (61%) | 8859 (51%) |
| 3+ | 33,944 (31%) | 3181 (41%) | 2329 (30%) | 7194 (41%) |
| On statin | 87,872 (81%) | 6679 (86%) | 5944 (77%) | 15,010 (86%) |
| On insulin | 4523 (4%) | 1352 (17%) | 144 (2%) | 1811 (10%) |

| | | | | |
|---|---|---|---|---|
| On ACE-inhibitor or ARB | 57,620 (53%) | 6394 (82%) | 4072 (52%) | 14,595 (83%) |
| Treatment arm | | | | |
| SGLT2-inhibitor | 39,991 (37%) | 3087 (40%) | 2852 (37%) | 7166 (41%) |
| DPP4-inhibitor/sulfonylurea | 68,438 (63%) | 4681 (60%) | 4916 (63%) | 10,369 (59%) |

* $pARR$ threshold 0.65%, matching treatment proportion as per treatment strategy based on uACR ≥3 mg/mmol.

Values for continuous variables are mean±SD or median (IQR); those for categorical variables are *n* (%)

Abbreviations: $pARR$ predicted absolute risk reduction, BMI body mass index, eGFR estimated glomerular filtration rate (calculated using the Chronic Kidney Disease Epidemiology Collaboration equation), uACR urinary albumin-creatinine ratio, ACE angiotensin-converting enzyme, ARB angiotensin-II receptor blocker, SGLT2-inhibitors sodium-glucose cotransporter-2 inhibitors, DPP4-inhibitors dipeptidyl peptidase-4 inhibitors.

**ESM Figure 1.** Flow diagram of study cohort inclusion and exclusion.

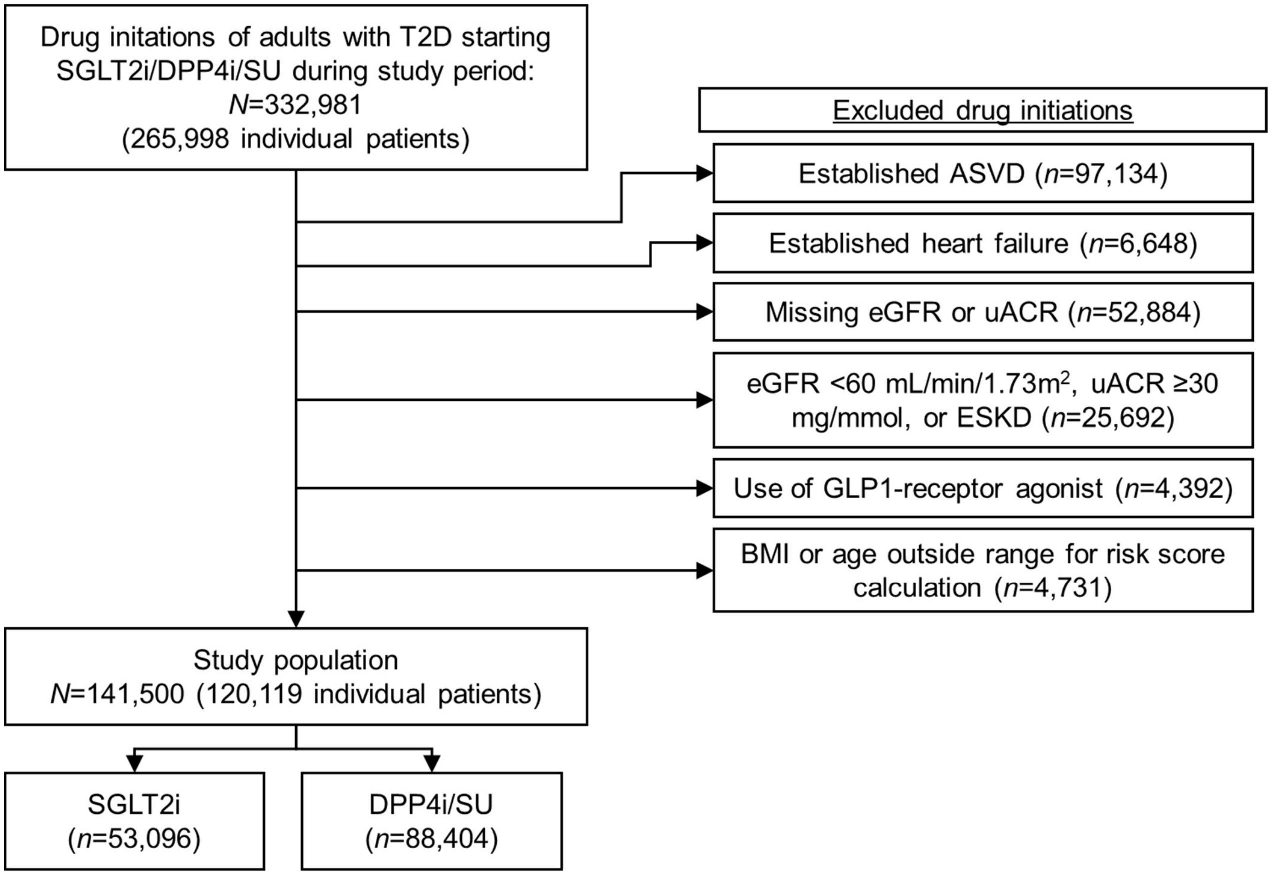


Abbreviations: T2D type 2 diabetes; SGLT2i SGLT2-inhibitors; DPP4i DPP4-inhibitors; SU sulfonylureas; ASVD atherosclerotic vascular disease (ischaemic heart disease or angina, peripheral vascular disease, revascularisation, stroke, or transient ischaemic attack); eGFR estimated glomerular filtration rate; uACR urinary albumin/creatinine ratio; ESKD end-stage kidney disease; BMI body mass index.

**ESM Figure 2.** Forest plot showing HRs with 95% confidence intervals for kidney disease progression (≥50% decline in eGFR, ESKD, or death due to kidney-related causes) over 3 years with SGLT2-inhibitors from trial meta-analysis (Nuffield Group, 2022) and estimated in this cohort using three analysis approaches. The analysis approaches used in this cohort included multivariable adjustment alone, doubly-robust overlap weighting, and doubly-robust inverse probability of treatment weighting (IPTW). The comparator arms were placebo in the meta-analysis of randomised controlled trials and DPP4-inhibitors/sulfonylureas in this study cohort. Follow-up time was mean 2.1 ±1.0 years in the SGLT2-inhibitor arm, and 2.2 ±1.0 years in the DPP4-inhibitor/sulfonylurea arm.

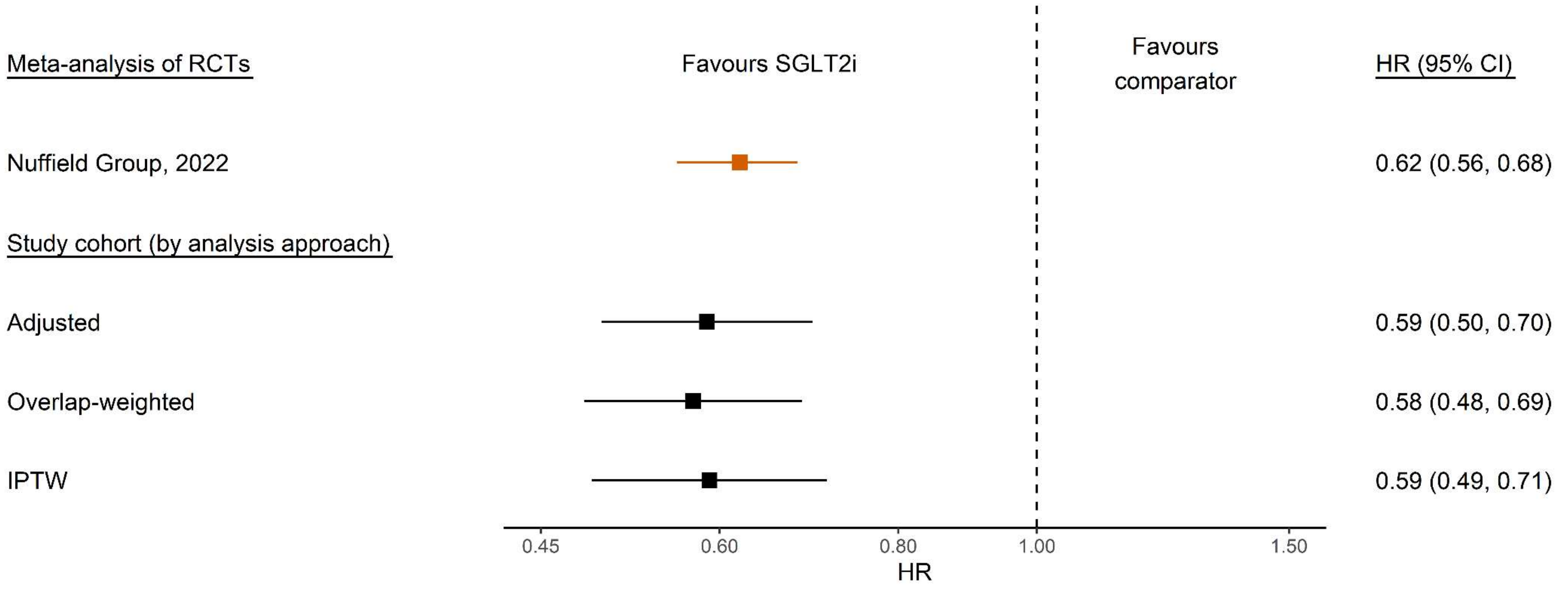


Abbreviations: RCT randomised controlled trial; SGLT2i SGLT2-inhibitor; CI confidence interval; IPTW inverse probability of treatment weighting

**ESM Figure 3.** Forest plot showing HRs with 95% confidence intervals for kidney disease progression (≥50% decline in eGFR, ESKD, or death due to kidney-related causes) over 3 years with pairwise comparisons between SGLT2-inhibitors, DPP4-inhibitors, and sulfonylureas, using 3 analytical approaches (multivariable adjustment, overlap weighting, and inverse probability of treatment weighting). In these analyses, individuals were also censored if starting a DPP4-inhibitor or sulfonylurea during follow-up whilst in a different treatment arm. They were subsequently included in the respective arm from the same date, allowing inclusion of two non-overlapping follow-up periods for these individuals (*n*=5270 drug episodes of starting a DPP4-inhibitor after an episode of starting a sulfonylurea or vice versa). Of note, in sensitivity analyses comparing SGLT2i-inhibitors to DPP4-inhibitors/sulfonylureas where only a single drug episode per participant was included, the association was similar (overlap-weighted HR 0.52, 95%CI 0.43, 0.61).

| SGLT2-inhibitors vs sulfonylureas | Events/subjects (SGLT2i) | Events/subjects (SU) | HR (95% CI) |
|---|---|---|---|
| Adjusted | 186/53,096 | 269/36,854 | 0.54 (0.43, 0.68) |
| Overlap-weighted | 186/53,096 | 269/36,854 | 0.53 (0.40, 0.70) |
| IPTW | 186/53,096 | 269/36,854 | 0.53 (0.40, 0.69) |

| SGLT2-inhibitors vs DPP4-inhibitors | Events/subjects (SGLT2i) | Events/subjects (DPP4i) | HR (95% CI) |
|---|---|---|---|
| Adjusted | 186/53,096 | 395/56,820 | 0.58 (0.48, 0.70) |
| Overlap-weighted | 186/53,096 | 395/56,820 | 0.61 (0.47, 0.79) |
| IPTW | 186/53,096 | 395/56,820 | 0.60 (0.48, 0.75) |

| DPP4-inhibitors vs sulfonylureas | Events/subjects (DPP4i) | Events/subjects (SU) | HR (95% CI) |
|---|---|---|---|
| Adjusted | 395/56,820 | 269/36,854 | 0.94 (0.79, 1.11) |
| Overlap-weighted | 395/56,820 | 269/36,854 | 0.86 (0.71, 1.05) |
| IPTW | 395/56,820 | 269/36,854 | 0.89 (0.73, 1.08) |

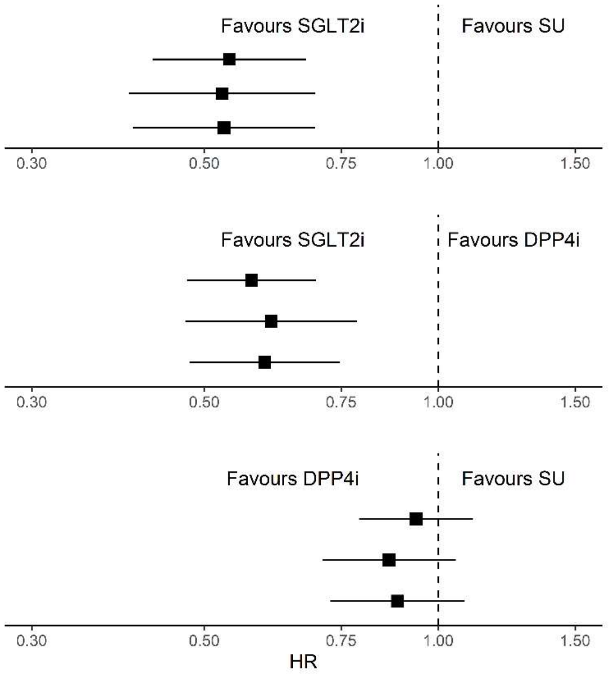


Abbreviations: SGLT2i SGLT2-inhibitors; DPP4i DPP4-inhibitors; SU sulfonylureas; CI confidence interval; IPTW inverse probability of treatment weighting.

**ESM Figure 4.** Calibration plot of predicted 3-year risk of kidney disease progression (≥50% decline in eGFR, ESKD, or death due to kidney-related causes) using the CKD-PC risk score compared with observed risk, in individuals treated with DPP4-inhibitors/sulfonylureas (*n*=88,404). Observed risk with 95% confidence intervals was calculated using Kaplan-Meier estimates per risk score decile.

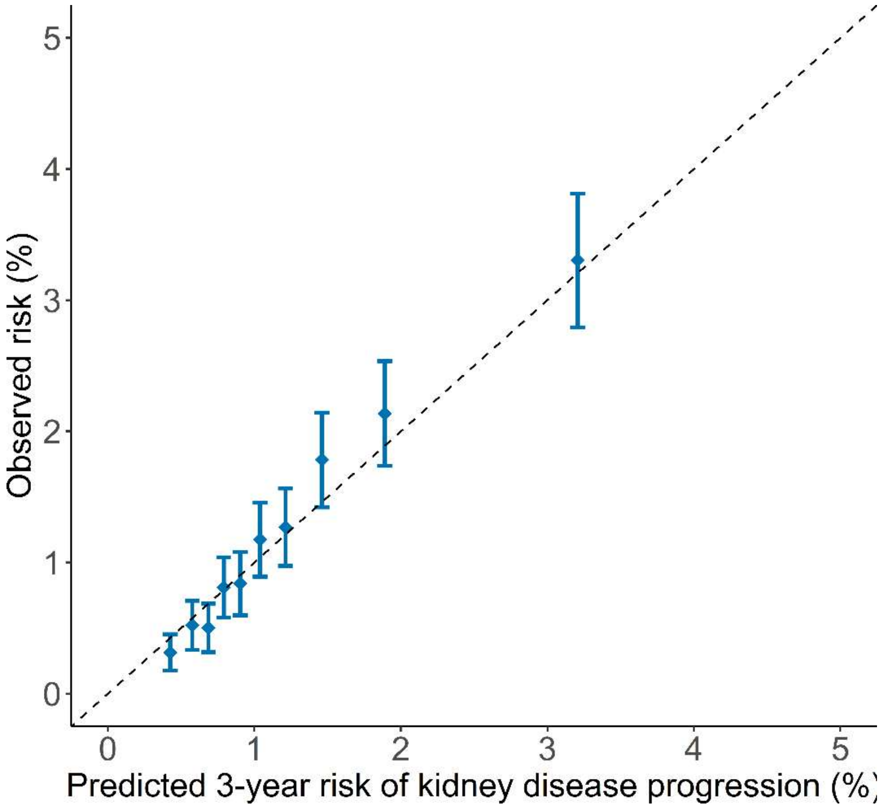


Abbreviations: CKD-PC Chronic Kidney Disease Prognosis Consortium.

**ESM Figure 5**. Calibration of model-predicted 3-year absolute risk reductions with SGLT2-inhibitor treatment (pARR) compared to counterfactual absolute risk reductions estimated from observed data using doubly robust overlap-weighted Cox models, using only one comparator drug at a time. Median and interquartile range are shown by deciles of pARR. Panel a shows calibration using DPP4-inhibitors as only comparator drug (calibration slope 1.14, 95% CI 1.13-1.16), and panel b shows calibration using sulfonylureas as only comparator drug (calibration slope 1.06, 95%CI 1.04-1.07).

a.

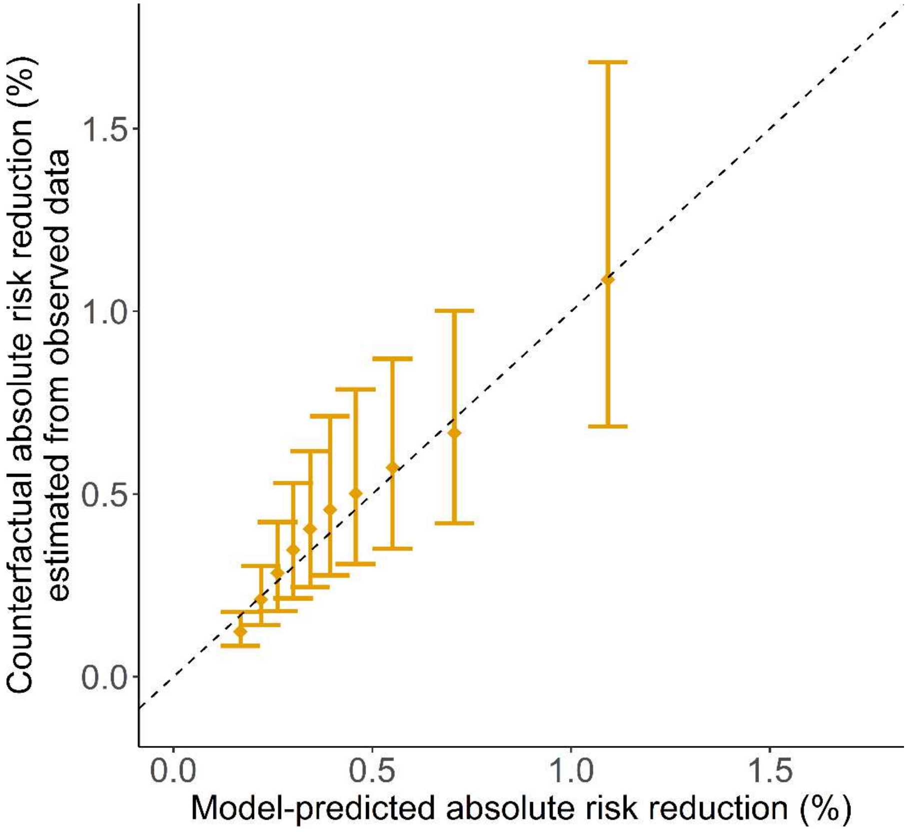

b.

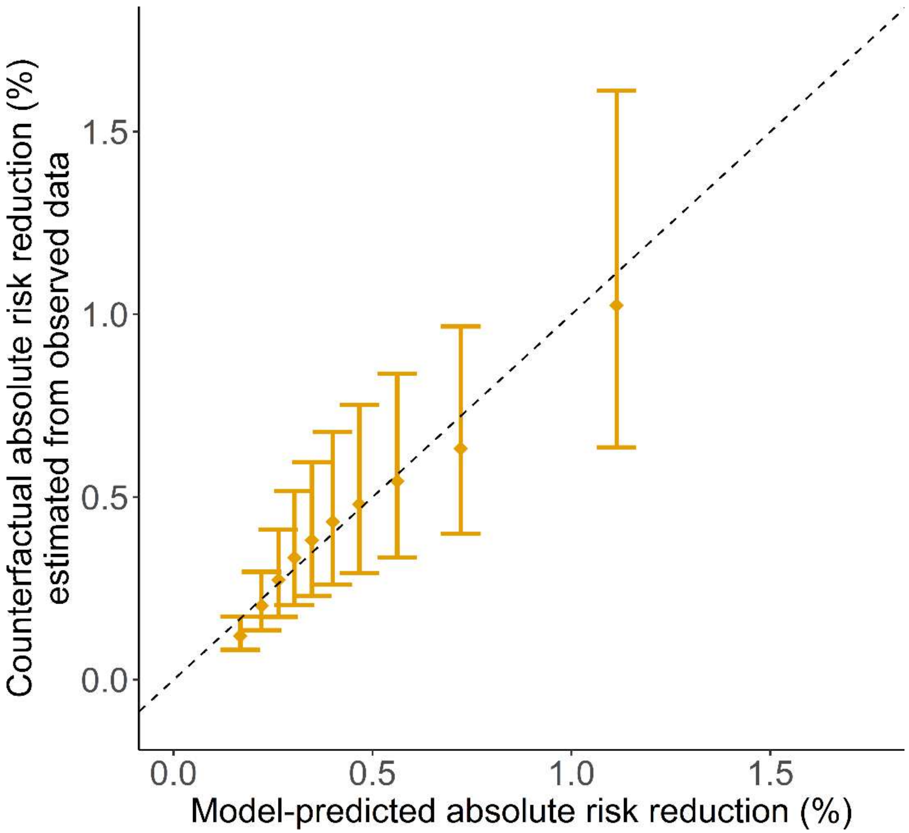
Counterfactual absolute risk reduction (%)
estimated from observed data
0.0
0.5
1.0
1.5
Model-predicted absolute risk reduction (%)

**ESM Figure 6.** Decision curve comparing net utility (also referred to as net benefit) of different treatment strategies for targeting individuals at risk of kidney disease progression (≥50% decline in eGFR, ESKD, or death due to kidney-related causes), by risk tolerance (also referred to as threshold probability; of 3-year risk of kidney disease progression). The solid blue line represents the treatment strategy based on uACR ≥3 mg/mmol (as recommended by current guidelines), the dashed orange line a treatment strategy based on $pARR$ ≥0.65%, where a comparable proportion of the population would be treated as under uACR ≥3 mg/mmol, and the solid yellow line represents a $pARR$-based strategy where the $pARR$ threshold varies according to the level of risk tolerance. The black solid line represents a strategy where everyone is treated, whereas the grey solid line represents a strategy where no one is treated. Higher net utility at a given level of risk tolerance indicates greater clinical utility of that strategy.

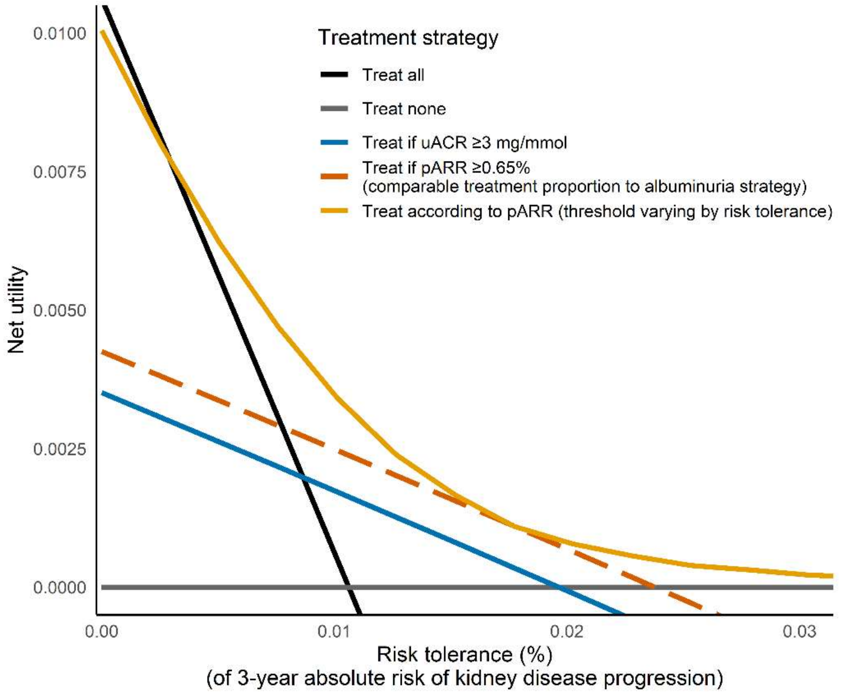


Abbreviations: pARR predicted absolute risk reduction with SGLT2-inhibitor treatment.

**ESM Figure 7.** Extended 5-year observational analyses of the impact of a treatment strategy based on $pARR$ ≥0.65% vs uACR ≥3 mg/mmol (currently recommended) on cumulative observed absolute risk reductions in kidney disease progression (≥50% decline in eGFR, ESKD, or death due to kidney-related causes). The ≥0.65% $pARR$ threshold was chosen to target a comparable proportion of the population as the ≥3 mg/mmol albuminuria threshold. Observed absolute risk reductions were derived from doubly robust overlap-weighted Cox proportional hazards models smoothed using a loess method.

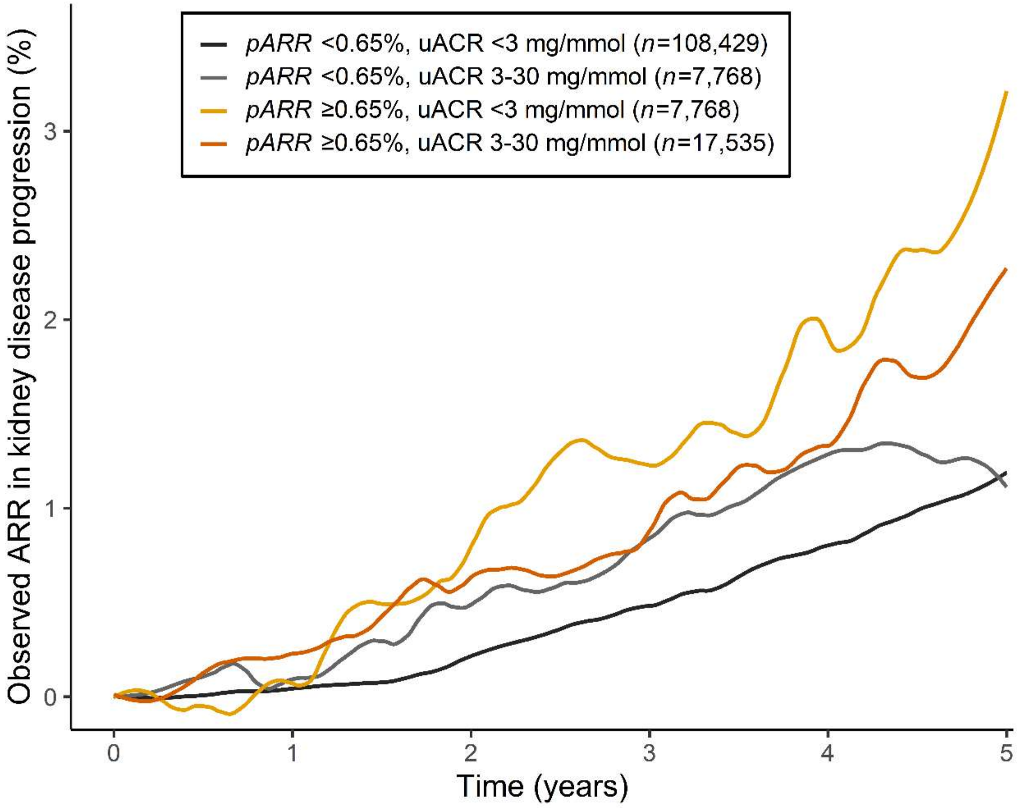


Abbreviations: ARR absolute risk reduction, pARR predicted absolute risk reduction with SGLT2-inhibitor treatment, uACR urinary albumin/creatinine ratio.

**ESM Figure 8.** Bar chart showing the observed 5-year absolute risk reduction in kidney disease progression (≥50% decline in eGFR, ESKD, or death due to kidney-related causes) with SGLT2-inhibitors in subgroups of individuals by treatment recommendation based on uACR ≥3 mg/mmol (striped vs solid) or predicted benefit ($pARR$; grey vs yellow). The ≥0.65% pARR threshold was chosen to target a comparable proportion of the population to the ≥3 mg/mmol albuminuria threshold. Observed absolute risk reductions were derived from doubly robust overlap-weighted Cox proportional hazards models smoothed using a loess method. Error bars depict 95% confidence intervals.

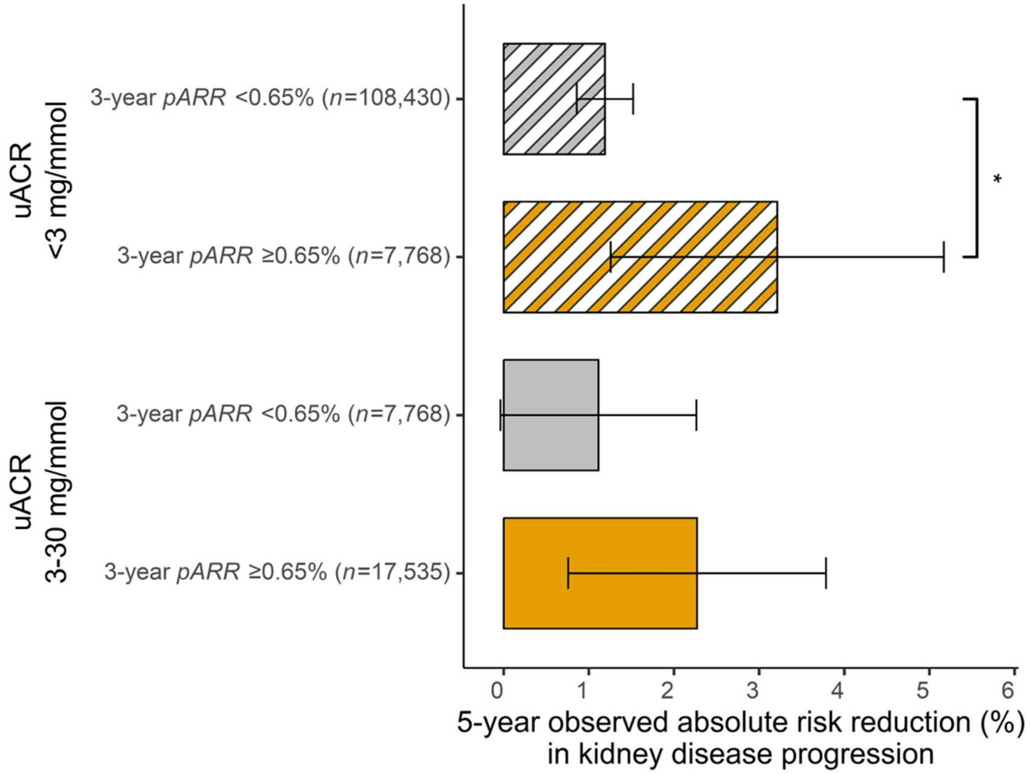


*p=0.05

**ESM Figure 9.** Forest plots showing HRs with 95% confidence intervals for secondary outcomes over 3 years with SGLT2-inhibitors compared to DPP4-inhibitors/sulfonylureas, stratified by uACR ≥3 mg/mmol (panel A) and $pARR$ ≥0.65% (panel B). Secondary outcomes included a composite outcome of 40% decline in eGFR, ESKD, or death due to kidney-related causes; the composite outcome of 50% decline in eGFR, ESKD, or death due to kidney-related causes (5 years rather than 3 years); progression to significant albuminuria (≥30 mg/mmol); diabetic keto-acidosis; amputation; and mycotic genital infection. Hazard ratios were derived from doubly robust overlap-weighted analyses. Baseline characteristics by $pARR$ are shown in ESM Table 4.

a.

| | Events/subjects (SGLT2i) | (DPP4i/SU) | HR (95% CI) |
|---|---|---|---|
| **40% eGFR decline / ESKD** | | | |
| uACR <3 mg/mmol | 357/43,078 | 1,039/73,119 | 0.68 (0.59, 0.79) |
| uACR 3-30 mg/mmol | 165/10,018 | 415/15,285 | 0.66 (0.54, 0.82) |
| **50% eGFR decline / ESKD (5 years)** | | | |
| uACR <3 mg/mmol | 208/43,078 | 837/73,119 | 0.53 (0.44, 0.63) |
| uACR 3-30 mg/mmol | 131/10,018 | 369/15,285 | 0.65 (0.52, 0.83) |
| **Progression to uACR ≥30 mg/mmol** | | | |
| uACR <3 mg/mmol | 23/43,078 | 64/73,119 | 0.83 (0.48, 1.43) |
| uACR 3-30 mg/mmol | 86/10,018 | 348/15,285 | 0.53 (0.40, 0.70) |
| **Diabetic keto-acidosis** | | | |
| uACR <3 mg/mmol | 178/43,078 | 155/73,119 | 1.80 (1.37, 2.36) |
| uACR 3-30 mg/mmol | 62/10,018 | 38/15,285 | 2.39 (1.41, 4.05) |
| **Amputation** | | | |
| uACR <3 mg/mmol | 51/43,078 | 64/73,119 | 1.26 (0.79, 1.99) |
| uACR 3-30 mg/mmol | 30/10,018 | 38/15,285 | 1.13 (0.64, 2.02) |
| **Mycotic genital infection** | | | |
| uACR <3 mg/mmol | 3,348/43,078 | 1,992/73,119 | 3.10 (2.90, 3.32) |
| uACR 3-30 mg/mmol | 709/10,018 | 451/15,285 | 2.76 (2.39, 3.19) |

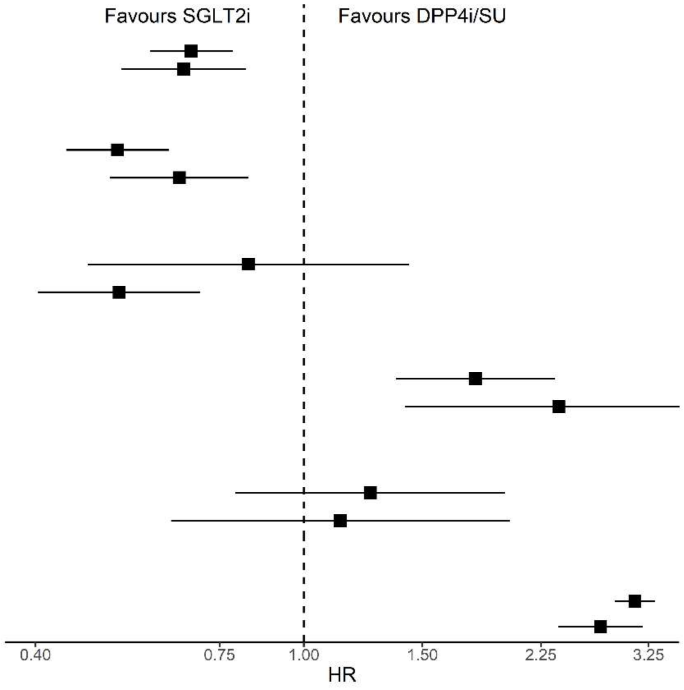

b.

| Outcome | Events/subjects (SGLT2i) | (DPP4i/SU) | HR (95% CI) |
|---|---|---|---|
| **40% eGFR decline / ESKD** | | | |
| pARR <0.65% | 326/42,843 | 934/73,355 | 0.69 (0.60, 0.80) |
| pARR ≥0.65% | 196/10,253 | 520/15,049 | 0.65 (0.54, 0.79) |
| **50% eGFR decline / ESKD (5 years)** | | | |
| pARR <0.65% | 182/42,843 | 755/73,355 | 0.52 (0.43, 0.63) |
| pARR ≥0.65% | 157/10,253 | 451/15,049 | 0.65 (0.52, 0.80) |
| **Progression to uACR ≥30 mg/mmol** | | | |
| pARR <0.65% | 35/42,843 | 125/73,355 | 0.61 (0.40, 0.93) |
| pARR ≥0.65% | 74/10,253 | 287/15,049 | 0.54 (0.40, 0.73) |
| **Diabetic keto-acidosis** | | | |
| pARR <0.65% | 168/42,843 | 141/73,355 | 2.13 (1.60, 2.84) |
| pARR ≥0.65% | 72/10,253 | 52/15,049 | 1.53 (0.99, 2.38) |
| **Amputation** | | | |
| pARR <0.65% | 43/42,843 | 65/73,355 | 1.12 (0.69, 1.80) |
| pARR ≥0.65% | 38/10,253 | 37/15,049 | 1.38 (0.79, 2.39) |
| **Mycotic genital infection** | | | |
| pARR <0.65% | 3,321/42,843 | 2,007/73,355 | 3.16 (2.95, 3.38) |
| pARR ≥0.65% | 736/10,253 | 436/15,049 | 2.55 (2.20, 2.95) |

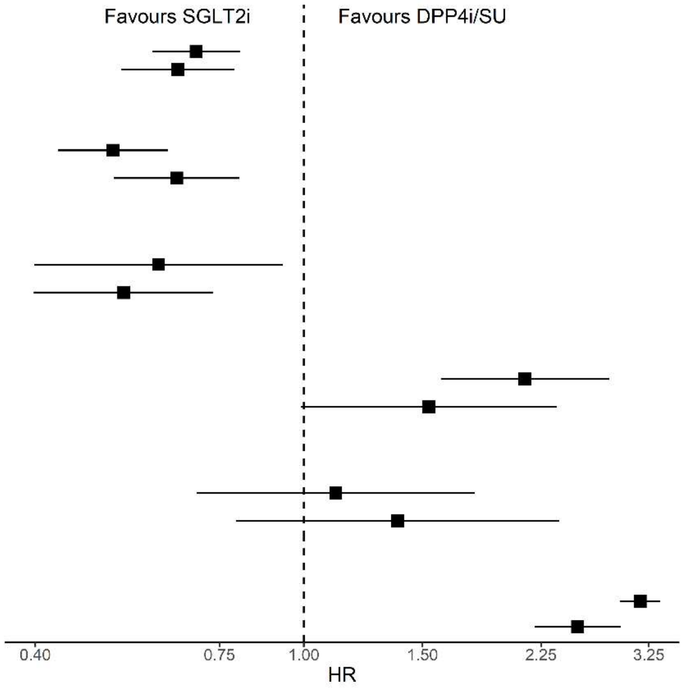


Abbreviations: SGLT2i SGLT2-inhibitors; DPP4i/SU DPP4-inhibitors/sulfonylureas; CI confidence interval; pARR predicted absolute risk reduction with SGLT2-inhibitor treatment.
